# Pulse pressure thresholds associated with cognitive impairment across diverse regional and ethnic populations

**DOI:** 10.64898/2026.05.26.26354116

**Authors:** Haoran Zhang, Richard N. Henson, Shihan Chen, Haoxuan Wen, Yijin Fang, Xuhao Zhao, Ting Pang, James B. Rowe, Xin Xu, Kamen A Tsvetanov

**Affiliations:** School of Public Health, the Second Affiliated Hospital of School of Medicine, Zhejiang University, Hangzhou 310058, China; Department of Clinical Neurosciences and Cambridge University Hopsitals NHS Trust, University of Cambridge, Cambridge CB2 0SZ, UK; Department of Psychiatry, University of Cambridge, Cambridge CB2 2QQ, UK; Department of Psychology, University of Cambridge, Cambridge CB2 3EB, UK; Medical Research Council Cognition and Brain Sciences Unit, Cambridge CB2 7EF, UK; Nanhu Brain-computer Interface Institute, Hangzhou 310058, China; MOE Frontier Science Center for Brain Science and Brain-Machine Integration, Zhejiang University School of Medicine, Hangzhou 310058, China

**Keywords:** Pulse Pressure, Thresholds, Cognition, Memory

## Abstract

**Background:** As dementia prevalence rises globally, it is critical to find preventions that target modifiable risk factors like blood pressure. Pulse pressure (PP), a marker of arterial stiffness, contributes independently to cognitive impairment. Yet, clinically interpretable thresholds for PP for cognitive decline remain undefined. We examined the independent association between PP and domain-specific cognitive trajectories and identified PP thresholds associated with greater cognitive decline across ethnically diverse regional populations.

**Methods:** Data were harmonized across three longitudinal cohorts (54,878 participants with up to 20 years follow-ups and 266,144 observations). Linear mixed-effects models identified a nonlinear association between PP and cognition (memory, orientation, and executive function), whereby cognitive decline accelerated after around 50 mmHg of pulse pressure, despite controlling for mean arterial pressure and dementia risk factors. Stratification based on PP thresholds (Low: PP <30; Normal: 30 to <50; Borderline: ≥50; and High: ≥60 mmHg), and tested for differences in memory decline across groups. Stratified analyses were similarly conducted across other blood pressure measures, racial, age and sex groups.

**Findings:** Non-linear associations indicated that memory decline was particularly noticeable for pulse pressure ≥60 mmHg. Compared with normal pulse pressure, ≥60 mmHg was associated with worse memory performance (pooled β −0.062 SD; 95% CI −0.107 to −0.016) and greater memory decline with age (−0.026 SD/year; −0.036 to −0.015), including among normotensive individuals. Findings were consistent across diverse regional cohorts (UK, US and China), racial groups, age strata and sexes.

**Interpretation:** Pulse pressure over 60 mmHg is associated with elevated cognitive risk, independent of blood pressure measures, even among normotensive individuals. These findings support pulse pressure thresholds as clinically interpretable and complementary markers of cognitive risk.

**Funding:** Alzheimer’s Society, NIHR Cambridge Biomedical Research Centre, NIHR BioResource, Wellcome Trust, Medical Research Council, Addenbrookes Charitable Trust, Natural Science Foundation of China

**Evidence before this study:** Blood pressure is recognized as a key modifiable target for dementia. However, previous findings indicate that systolic blood pressure alone may not fully capture vascular aging contributions to cognitive decline. Pulse pressure - the difference between systolic and diastolic blood pressure - provides insights into the pulsatile component beyond conventional indices. Prior studies have reported associations between pulse pressure and cognitive impairment across individuals, but few have examined domain-specific impairment (e.g., memory) across diverse ethnoregional populations over extended periods of ageing within individuals. Evidence from arterial stiffness and pulsatile biology suggests that cognitive risk may increase non-linearly, accelerating only beyond specific blood pressure levels. Defining target thresholds is crucial for guiding public health and clinical interventions, yet to date, no clinically interpretable pulse pressure threshold has been established.

**Added value of this study:** We demonstrate for the first time that clinically interpretable pulse pressure thresholds predict cognitive impairment independent of traditional blood pressure measures. By harmonizing three ethnically diverse regional populations (United Kingdom, United States and China), this study confirmed an overall association between higher pulse pressure and poorer cognitive function. We further showed that this association is non-linear, was most evident in memory function, and was independent of traditional blood pressure measures and established modifiable dementia risk factors. Pulse pressure ≥60 mmHg was associated with worse memory performance and greater memory decline, including among normotensive individuals, with consistent findings across observed across multiple cohorts, racial groups (White, Black and Asian), age strata (mid-life and late-life) and both sexes. Furthermore, participants with pulse pressure ≥50 mmHg had greater memory decline even among normotensive individuals.

**Implications of all the available evidence:** Our findings support the use of pulse pressure thresholds above 60 mmHg as a clinically interpretable and complementary marker to systolic blood pressure for assessing dementia risk across diverse populations, even among normotensive individuals. Timely monitoring pulse pressure above 50 mmHg may facilitate early identification of individuals at risk for cognitive decline.

## 1 Introduction

Global population ageing has resulted in an increasing burden of cognitive decline and dementia, despite progress in addressing several risk factors.^1,2^ The number of people of living with dementia worldwide is projected to rise substantially, particularly in lower-income countries, reaching an estimated 153 million by 2050.^3^ Identifying the modifiable risk factors for cognitive decline is therefore a public health priority for preventing or delaying dementia.^3,4^

Midlife hypertension has been identified as a modifiable risk factor of cognitive decline and dementia later in life.^3,5,6^ However, evidence on the association between blood pressure (BP) and cognitive outcomes remains inconsistent, with mixed findings from studies of systolic blood pressure (SBP) (based on, for example, the clinical definition of hypertension as SBP above 140mmHg).^6,7^ This is likely due to SBP alone not fully capturing arterial stiffness and heterogeneity in population characteristics. Consistent with this, SBP interventions showed little if any benefit for cognition, suggesting that there is another mechanism through which blood pressure contributes to cognitive impairment and dementia.^7–9^

Another way of looking at BP is through two components: a steady state component – the average of systolic and diastolic pressures, e.g. “mean arterial pressure” (MAP) – and a pulsatile component – the difference between systolic and diastolic, termed “pulse pressure” (PP).^10^ PP is thought to reflect arterial stiffness and increased transmission of pulsatile energy into the vascular tree, reaching cerebrovascular arterioles and capillaries, and potentially leading to impaired cerebral blood flow and hypoxia.^10–13^ Recent studies have demonstrated associations between PP and cognitive impairment in mid- and late-life adults, providing information beyond traditional blood pressure measures (e.g., SBP and MAP).^4,11,14^ However, most studies have focused on single populations and global cognition, with limited attention to domain-specific outcomes (e.g., memory) or ethnic and regional differences.^4,6,15^ In addition, as arteries lose elasticity and stiffen with increasing age, age-specific variation in the association between PP and cognition needs to be considered.^6,12^ Therefore, the present study aims to systematically examine whether PP is independently associated with cognitive function over extended periods of ageing.

As suggested by arterial stiffness and pulsatile biology, cognitive risk may increase non-linearly, accelerating only above certain PP levels.^13^ Identifying clinically relevant PP thresholds is therefore important for public health and clinical strategies, such as informing screening, prevention, and therapeutic decision-making.^7,11^ Establishing such thresholds may enable earlier identification of individuals at elevated cognitive risk; individuals who would not be detected using conventional BP measures alone. Prior studies largely examined PP as a continuous composite measure, rather than identifying clinically interpretable thresholds.^14–17^ Guidelines from the European Society of Hypertension state that, in older adults, a PP exceeding 60 mmHg is indicative of increased cardiovascular risk.^18,19^ To our knowledge, no study has explored optimal PP thresholds f for domain-specific cognitive outcomes. Therefore, using large, diverse cohorts with harmonised cognitive assessments, the present study aimed to establish PP thresholds predictive of cognitive impairment independent of traditional blood pressure measures.

## 2 Methods

### 2-1 Participants

The present study leveraged individual-level data from three nationally-representative longitudinal cohorts: the Health and Retirement Study (HRS) from the United States^20^, the English Longitudinal Study of Ageing (ELSA) from the United Kingdom^21^, and the China Health Retirement Longitudinal Study (CHARLS) from China^22^. Data from wave 8 (2006) to wave 15 (2020) in HRS, from wave 0 (1998) to wave 11 (2022) in ELSA, and from wave 0 (2011) to wave 4 (2020) in CHARLS were used. Figure 1 illustrates the analytical approach. Given that these cohorts were continuously refreshed with newly recruited participants across waves, and to maximize the sample size and enhance the generalization of results, we included all available participants, defining baseline on an individual basis, as the wave in which a participant first had an available BP measurement, relabelled as wave 0 (Figure 2).

**Figure 1.**
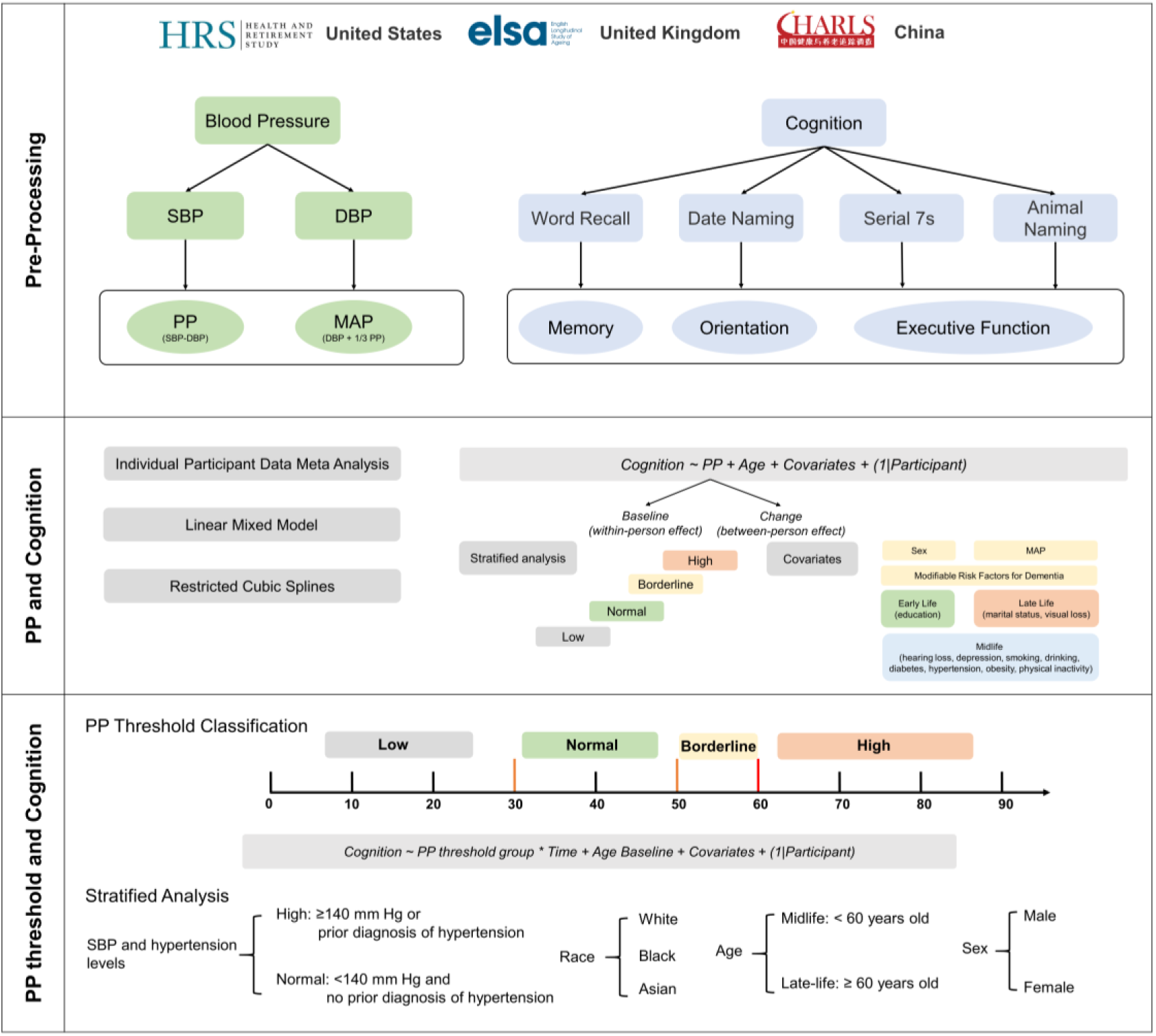
A schematic representation of the main stages in the data processing and analysis pipeline CHARLS=China Health and Retirement Longitudinal Study. DBP=diastolic blood pressure. ELSA=English Longitudinal Study HRS=Health and Retirement Study. MAP=mean arterial pressure. PP=pulse pressure. SBP=systolic blood pressure.

**Figure 2.**
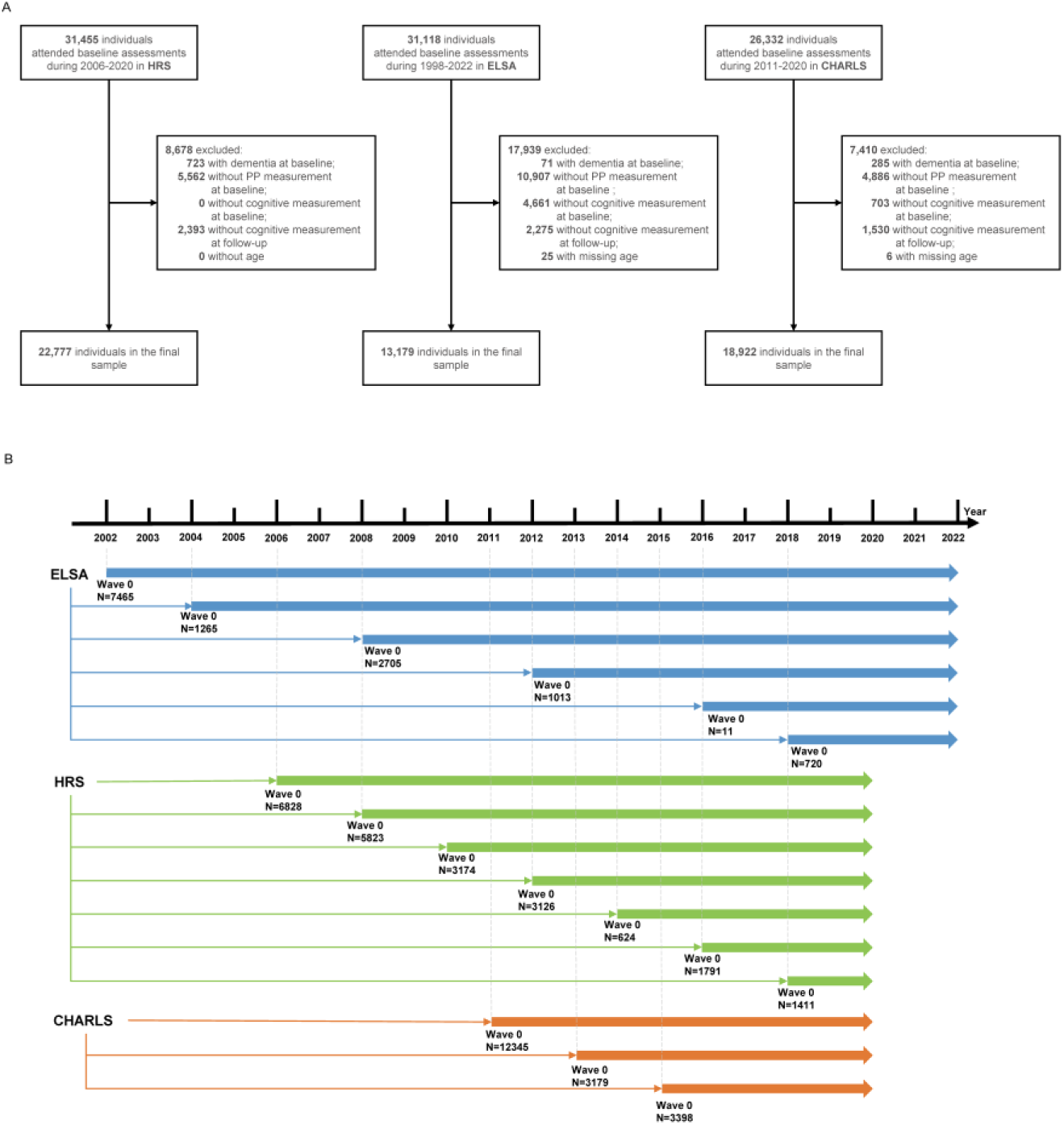
A. Study Enrolment and B. Newly recruited sample sizes and age ranges across waves in each study CHARLS=China Health and Retirement Longitudinal Study. ELSA=English Longitudinal Study of Ageing. HRS=Health and Retirement Study. PP=pulse pressure.

Participants were excluded if they met any of the following criteria: (1) diagnosis of dementia or memory problems at baseline, (2) absence of PP measurement at baseline, (3) absence of all cognitive measurements at baseline, (4) absence of all cognitive measurements during the follow-up, and (5) missing age information.

All study participants signed informed consent before the study. The HRS was approved by the Institutional Review Board at the University of Michigan and the National Institute on Aging (HUM00061128). ELSA was approved by the London Multicentre Research Ethics Committee (MREC/01/2/91). CHARLS was approved by the Peking University Institutional Review Board (IRB00001052–11015).

### 2-2 Pulse Pressure

We used BP measurements available in waves 0 to 6 from HRS, in waves 0, 2, 4, 6, 8 and 9 from ELSA, and in wave 0, 1 and 2 from CHARLS. In the ELSA, blood pressure at wave 0 was used as the baseline blood pressure for wave 1, since no BP measurement was conducted at wave 1.^23^ For consistency with other studies, wave 1 in the ELSA was then relabelled as wave 0.

Resting SBP and DBP were assessed. They were then used to calculate PP (SBP−DBP) and MAP (1/3 SBP + 2/3 DBP).^24^ BP were taken three times each. If there were three valid BP measurements, we reported the mean of the second and third measurements. Else if there were one or two valid measurements, we report the mean of all valid measurements. To distinguish within-person from between-person effects in the longitudinal data, PP was further decomposed into baseline PP and PP change, both of which were mean-centered. PP change was calculated as the difference between PP at each wave and PP at baseline.

PP thresholds were informed by prior theory and literature, with 50 and 60 mmHg indicating potential cardiovascular risk through mechanical pulsatile stress and 30 mmHg suggesting possible blood loss.^13,18,25–28^ Participants were classified into four groups based on a previously-established approach:^23^ 1) Low, defined as having PP values below 50 mmHg in all waves, with at least one wave below 30 mmHg; (2) Normal, defined as PP values consistently ranging from 30 to less than 50 mmHg across all measurement waves; (3) Borderline, defined as participants who did not meet the criteria for the Low or Normal groups and had PP values below 60 mmHg across all measurement waves; and (4) High, defined as having at least one measurement wave with a PP value of 60 mmHg or higher. Detailed definitions of PP categories were shown in Table S1. This approach reflects whether individuals successfully maintain their PP below a specific threshold over time.

### 2-3 Cognitive assessments

Three domains of cognitive assessments - memory, orientation and executive function - were conducted across all waves of studies. Memory was evaluated by immediate and delayed recall of ten words.^16,23^ Orientation was evaluated by date-naming tests.^16,23,29^ For executive function, serial-sevens tests were implemented in CHARLS and HRS,^23,29^ while the animal-naming fluency test was implemented in ELSA.^16^ Detailed definitions of cognitive assessments were shown in Table S2. To create comparable cognitive measures across studies and provide stable estimates in the pooled analysis, z scores were generated by standardizing to baseline (i.e., each domain test score was subtracted by the mean and then divided by the standard deviation (SD) of the baseline domain scores), with higher scores indicating better cognitive performance.^16,23,30^ Cognitive z scores were winsorized at ±3 standard deviations from the mean.^31^

### 2-4 Covariates

Covariates included age, sex, MAP and 11 modifiable factors of dementia available in the datasets. The modifiable factors were based on previously established criteria and covered early life (less education), midlife (hearing loss, depression, physical inactivity, diabetes, smoking, hypertension, obesity, excessive alcohol consumption), and late-life (marital status, and visual loss).^3^ Detailed definitions of modifiable factors of dementia are shown in Table S3. Missing covariates were imputed using multiple imputation by chained equations.^32^

### 2-5 Statistical Analysis

Demographics characteristics were compared among studies. Categorical variables were expressed as frequencies and percentages, and continuous variables were expressed as mean and SD. Chi-square test and Kruskal-Wallis were used for categorical and continuous variables, respectively.

#### Data-driven, nonlinear effects of PP on Cognition

Linear mixed models and restricted cubic splines were used to explore the linear and nonlinear association between PP and cognition.^33^ To better capture longitudinal effects, we decomposed PP into a between-person component (baseline PP) and a within-person component (change in PP over time), reflecting established approaches for separating cross-sectional differences from longitudinal change.^34^ We hypothesised that the effect of PP change on cognition vary across levels of baseline PP, consistent with vascular ageing and arterial stiffness mechanisms.^34^ Based on these considerations, and supported by exploratory analyses presented in the Supplementary Method, we fitted a parsimonious model in which between-person component, namely baseline PP, was modelled non-linearly, while the within-person component, i.e. PP change, was specified as a linear effect. This specification had higher marginal R2 (Table S8), and facilitated interpretation of changes, particularly given the relatively short follow-up period (2–4-years), during which within-person changes in PP are typically modest at the population level^35^. The model is specified using Wilkinson’s notation as follows:^36^

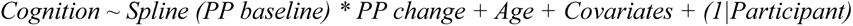

The optimal number of knots for the restricted cubic splines was evaluated by fitting models with three to five knots. Covariates included sex, MAP, early, midlife, and late-life of modifiable factors.

#### Threshold effects of PP on cognition

To facilitate interpretation of the nonlinear associations, additional stratified analyses were performed to assess the associations between PP and cognition within the pre-defined PP threshold groups. To assess the impact of these groups on trajectories of cognitive decline, the association between PP threshold group and cognitive decline (SD/year) during follow-up was evaluated by linear mixed models including PP threshold group, time (duration since baseline), group × time, age at baseline, and the covariates mentioned above:

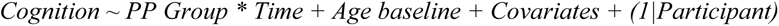

The effects of interest were the main effect of PP Group, and the interaction between PP group and time, which would indicate that rates of cognitive decline differ by PP group. As participants were enrolled at different baseline waves within each cohort, the effect of sub-cohorts and number of visits were also considered and included as a fixed effects to account for cohort effects and differences in visit patterns.^37^ Two-step individual participant data (IPD) meta-analyses with random effects were employed to pool effects across studies. *I*^2^ and τ^2^ statistics were reported to reflect heterogeneity between studies.^38^ Finally, further analyses were performed stratified by SBP levels, by race (White, Black, and Asian), by age (midlife: <60 and late-life: ≥60 years old), and by sex. Normal SBP was defined as SBP <140 mmHg at all waves and no prior diagnosis of hypertension; and High SBP was defined as SBP ≥140 mmHg at least at one wave or prior diagnosis of hypertension.^23^

Several sensitivity analyses were also conducted. Firstly, the nonlinear effect of baseline age was added to the models. Secondly, to address the potential white coat effect, which may lead to transient elevation of PP during measurement, we further distinguished participants with a single PP wave exceeding the threshold from those with more than one wave exceeding the threshold, i.e. addressing transient exposure versus persistent exposure effects. Thirdly, to account for treatment status (treated vs untreated hypertension), antihypertensive medication use (yes/no) was included as a covariate. Fourthly, to mitigate potential bias in the estimation of the baseline intercept arising from including individuals with only a single PP measurement, participants without follow-up PP assessments were excluded. We also applied stratified analysis to account for differences in visiting patterns,^37^ defining two groups according to number of PP visits: ≤2 versus ≥3 visits. Fifthly, participants with baseline stroke and heart disease were excluded to reduce potential confounding from cardiovascular conditions. Sixthly, analyses were repeated using complete-case datasets, and the results were compared with those obtained from the imputed datasets.

Lastly, we pooled data across studies and conducted one-step IPD meta-analyses to reassess the results.^39^ All analyses were conducted using R version 4·4·1 (R Project for Statistical Computing). Statistical significance was defined as a 2-sided p < 0.05. Effect sizes are reported in terms of Betas. The R code is publicly available at: https://github.com/haoranzhang0624-lang/PP-Threshold-And-Cognition.git.

## 3 Results

### 3-1 Baseline characteristics of participants

The present study included 54,878 participants, comprising a total of 266,144 observations (Figure 2, Table 1). Specifically, we included 22,777 HRS participants (maximum wave: 8; mean age: 63.6±10.7 years; 41.6% male), 13,179 ELSA participants (maximum wave: 11; mean age: 62.7±9.6 years; 44.4% male), and 18,922 CHARLS participants (maximum wave: 5; mean age: 57.4±9.8 years; 47.0% male). Rate of missing data varied between 0% and 33.7% (Table S4). Characteristics of the participants across different recruitment waves within each study were outlined in Table S5-7.

**Table 1:**
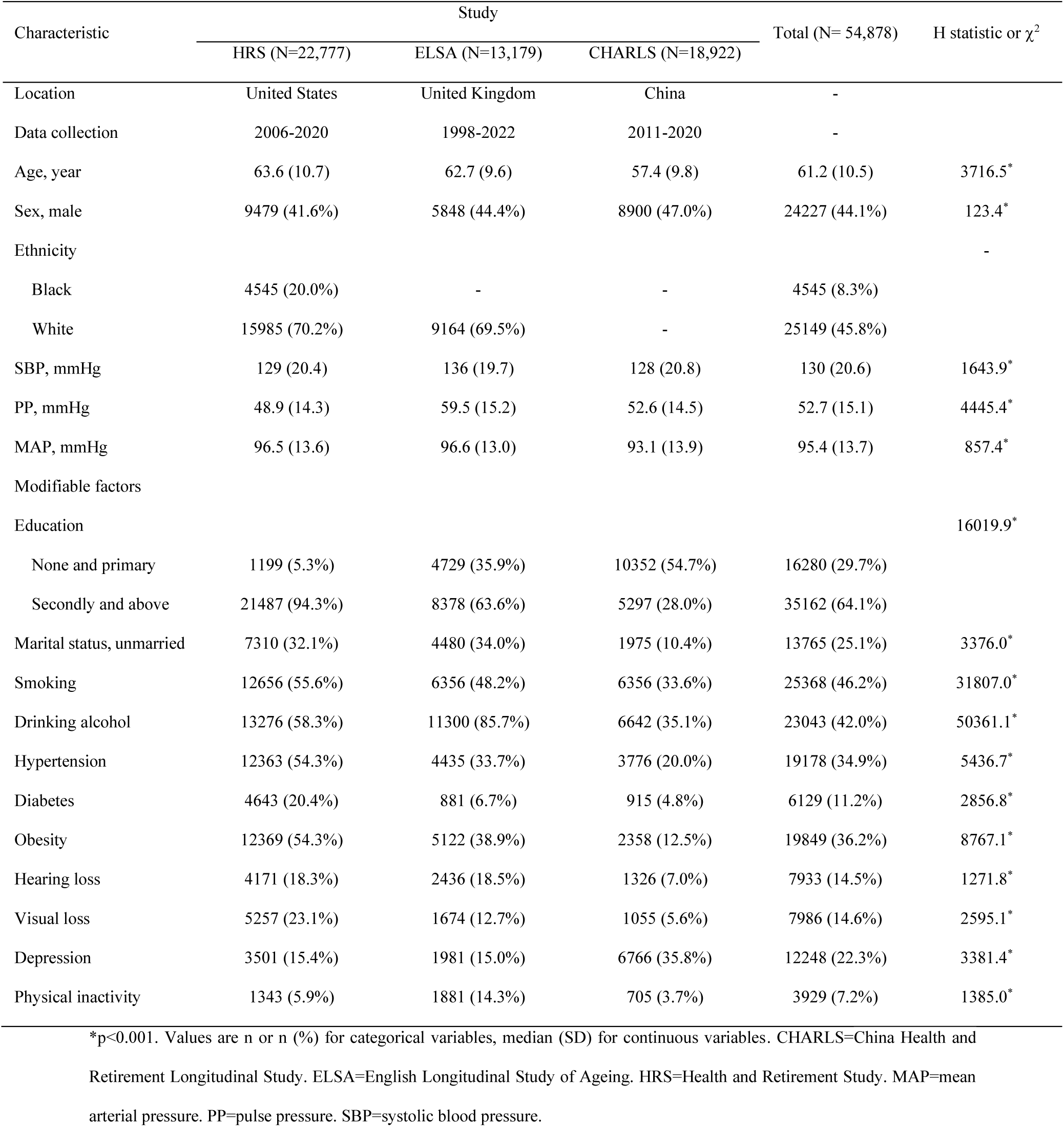
Baseline characteristics of participants in all cohorts.

| Characteristic | Study | | | Total (N= 54,878) | H statistic or $\chi^2$ |
| --- | --- | --- | --- | --- | --- |
|  | HRS (N=22,777) | ELSA (N=13,179) | CHARLS (N=18,922) |  |  |
| Location | United States | United Kingdom | China | - |  |
| Data collection | 2006-2020 | 1998-2022 | 2011-2020 | - |  |
| Age, year | 63.6 (10.7) | 62.7 (9.6) | 57.4 (9.8) | 61.2 (10.5) | 3716.5* |
| Sex, male | 9479 (41.6%) | 5848 (44.4%) | 8900 (47.0%) | 24227 (44.1%) | 123.4* |
| Ethnicity |  |  |  |  | - |
| Black | 4545 (20.0%) | - | - | 4545 (8.3%) |  |
| White | 15985 (70.2%) | 9164 (69.5%) | - | 25149 (45.8%) |  |
| SBP, mmHg | 129 (20.4) | 136 (19.7) | 128 (20.8) | 130 (20.6) | 1643.9* |
| PP, mmHg | 48.9 (14.3) | 59.5 (15.2) | 52.6 (14.5) | 52.7 (15.1) | 4445.4* |
| MAP, mmHg | 96.5 (13.6) | 96.6 (13.0) | 93.1 (13.9) | 95.4 (13.7) | 857.4* |
| Modifiable factors |  |  |  |  |  |
| Education |  |  |  |  | 16019.9* |
| None and primary | 1199 (5.3%) | 4729 (35.9%) | 10352 (54.7%) | 16280 (29.7%) |  |
| Secondly and above | 21487 (94.3%) | 8378 (63.6%) | 5297 (28.0%) | 35162 (64.1%) |  |
| Marital status, unmarried | 7310 (32.1%) | 4480 (34.0%) | 1975 (10.4%) | 13765 (25.1%) | 3376.0* |
| Smoking | 12656 (55.6%) | 6356 (48.2%) | 6356 (33.6%) | 25368 (46.2%) | 31807.0* |
| Drinking alcohol | 13276 (58.3%) | 11300 (85.7%) | 6642 (35.1%) | 23043 (42.0%) | 50361.1* |
| Hypertension | 12363 (54.3%) | 4435 (33.7%) | 3776 (20.0%) | 19178 (34.9%) | 5436.7* |
| Diabetes | 4643 (20.4%) | 881 (6.7%) | 915 (4.8%) | 6129 (11.2%) | 2856.8* |
| Obesity | 12369 (54.3%) | 5122 (38.9%) | 2358 (12.5%) | 19849 (36.2%) | 8767.1* |
| Hearing loss | 4171 (18.3%) | 2436 (18.5%) | 1326 (7.0%) | 7933 (14.5%) | 1271.8* |
| Visual loss | 5257 (23.1%) | 1674 (12.7%) | 1055 (5.6%) | 7986 (14.6%) | 2595.1* |
| Depression | 3501 (15.4%) | 1981 (15.0%) | 6766 (35.8%) | 12248 (22.3%) | 3381.4* |
| Physical inactivity | 1343 (5.9%) | 1881 (14.3%) | 705 (3.7%) | 3929 (7.2%) | 1385.0* |

\*p<0.001. Values are n or n (%) for categorical variables, median (SD) for continuous variables. CHARLS=China Health and Retirement Longitudinal Study. ELSA=English Longitudinal Study of Ageing. HRS=Health and Retirement Study. MAP=mean arterial pressure. PP=pulse pressure. SBP=systolic blood pressure.

### 3-2 Nonlinear association of PP with Cognition

A significant nonlinear association between PP and memory was found, even after adjusting for covariates (Figure 3). By decomposing PP into between-person (baseline) and within-person (change) components, a significant interaction confirmed that the effect of PP change differed according to baseline PP (see Tables S8-S9 for model comparisons). As shown by the blue lines in Figure 3, at lower baseline PP levels (approximately 20-30mmHg), overall memory performance was lower, but increases in PP change were positively associated with memory. In the mid-range of baseline PP (approximately 30-50mmHg), where performance was the highest (peaking around 40mmHg), no significant association between PP change and memory was observed. At higher baseline PP levels (>50mmHg), memory performance declined again, and increases in PP were significantly associated with poorer memory, i.e. the within-participant slope became negative.

**Figure 3:**
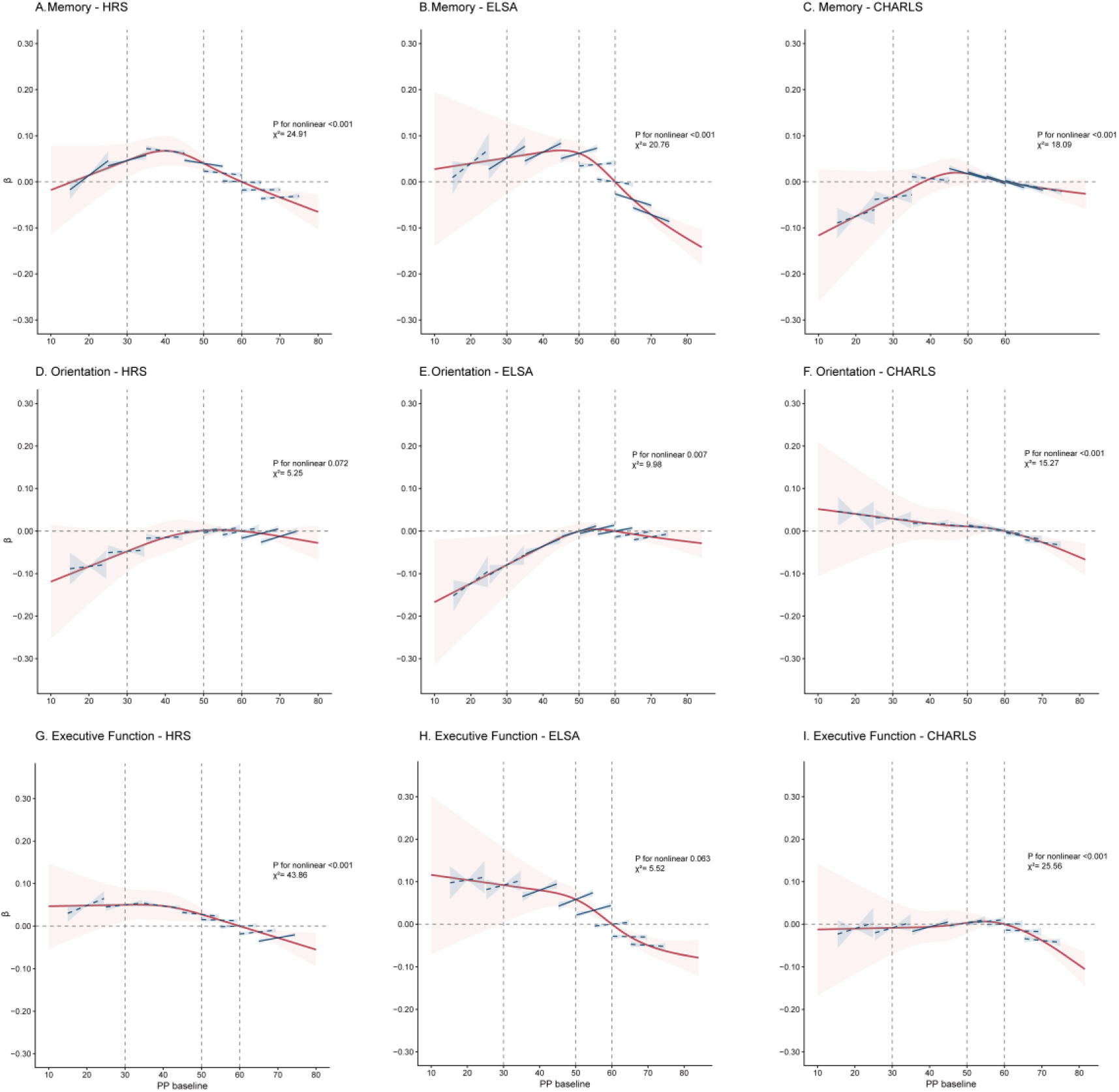
Associations of PP baseline and change with memory (A-C), orientation (D-F), and execution (G-I) in HRS (A, D, G), ELSA (B, E, H), and CHARLS (C, F, I). Models were specified as: Cognition ∼ Spline (PP baseline) * PP change + Age + Covariates + (1|Participant). P values represents the overall test for nonlinearity estimated from linear mixed-effects models. The red line represents the coefficients and confidence intervals of PP baseline (reference PP baseline=60 mmHg), whereas the blue line represents the coefficients and confidence intervals of PP change at the corresponding level of PP baseline (reference PP change=0 mmHg). PP change is presented within the range of −5 to +5, with solid lines indicating statistically significant associations and dashed lines indicating non-significant associations. Nonlinear effects of baseline PP were modelled using a restricted cubic spline with four knots. Covariates included sex, mean arterial pressure, early life factor (education), midlife factors (hearing loss, depression, smoking, drinking, diabetes, hypertension, obesity, and physical inactivity), and late life factors (marital status and visual loss). CHARLS=China Health and Retirement Longitudinal Study. ELSA=English Longitudinal Study of Ageing. HRS=Health and Retirement Study. PP=pulse pressure.

Although non-linear patterns were also observed for orientation and executive function in some cohorts, these associations were not so consistently replicated (Figure S2-3, D-I). Accordingly, subsequent analyses focus primarily on memory.

### 3.3 PP thresholds associated with cognition

To interpret the nonlinear relationship between PP and memory, and check the validity of the pre-specified PP thresholds, we stratified further analyses by PP group, and tested differences in baseline memory, as well as rate of decline of memory (see Table S1). Characteristics of the participants across four PP groups within each study are shown in Table S10-12.

As shown in Table 2, compared with individuals with Normal BP, those with High PP experienced significant overall memory impairment (pooled β = −0.062 SD; 95% CI: −0.0107 to −0.016) as well as greater memory decline (pooled β = −0.026 SD/year; 95% CI: −0.036, −0.015). Notably, the Borderline PP group (pooled β = −0.010 SD/year; 95% CI: −0.018, −0.003) also showed a greater decline in memory scores than the Normal group. The Low PP group did not differ significantly from the Normal group.

**Table 2:**
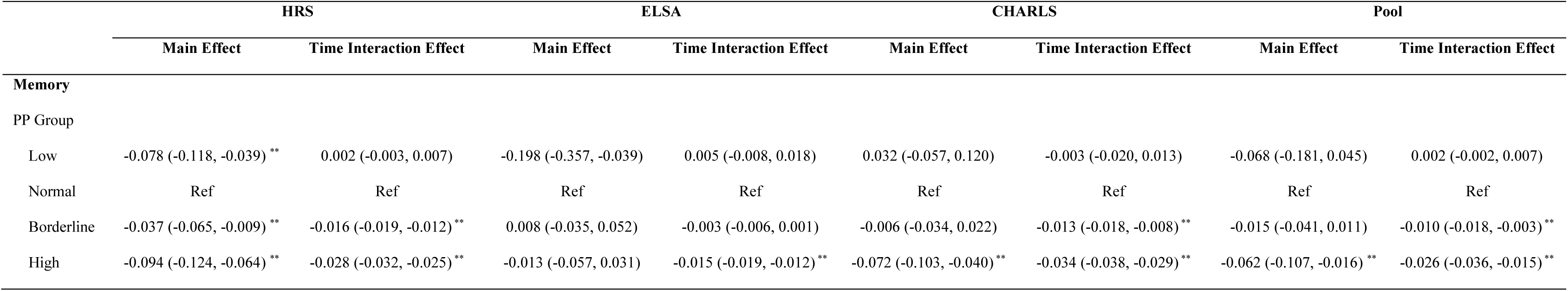
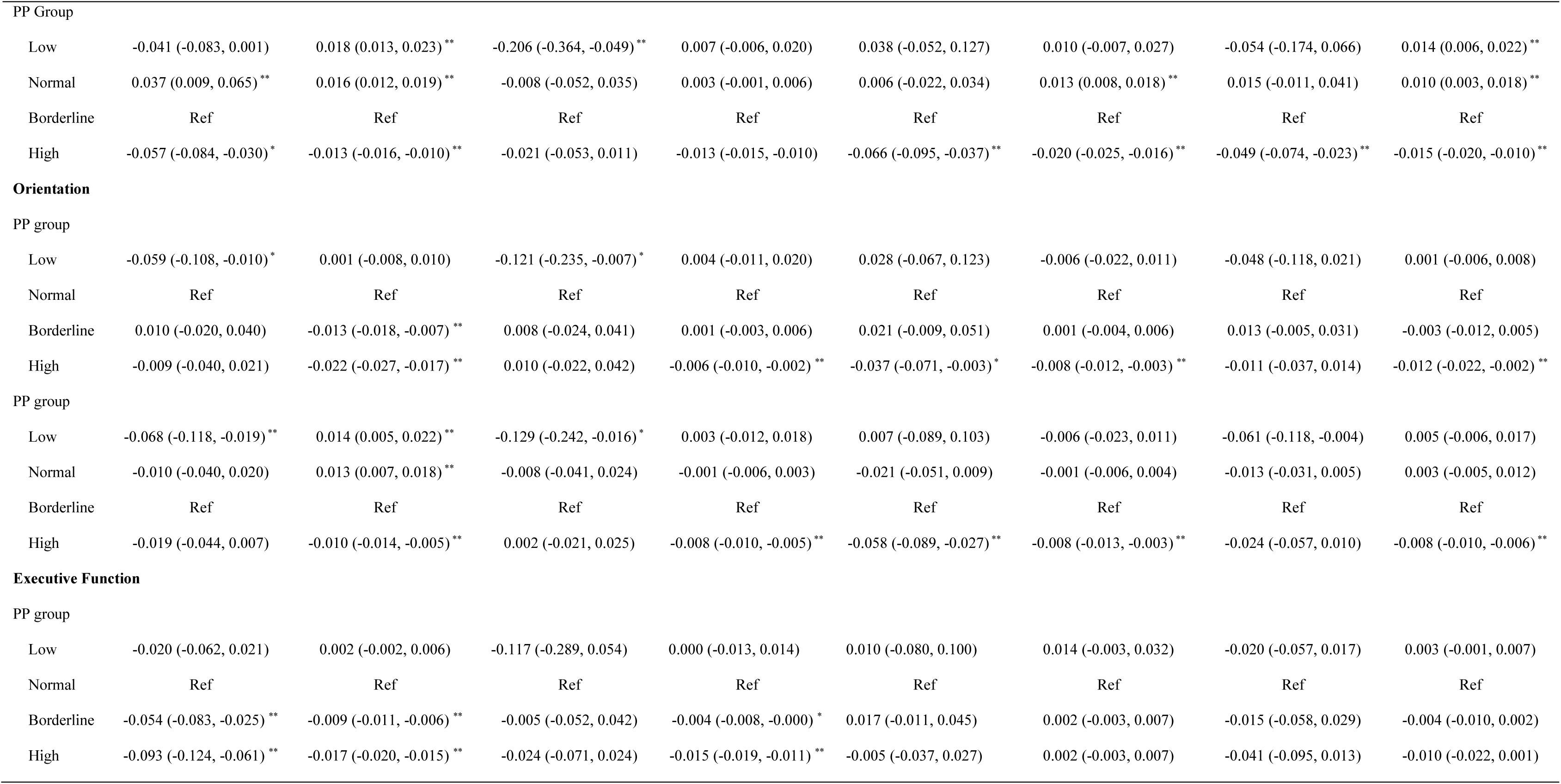
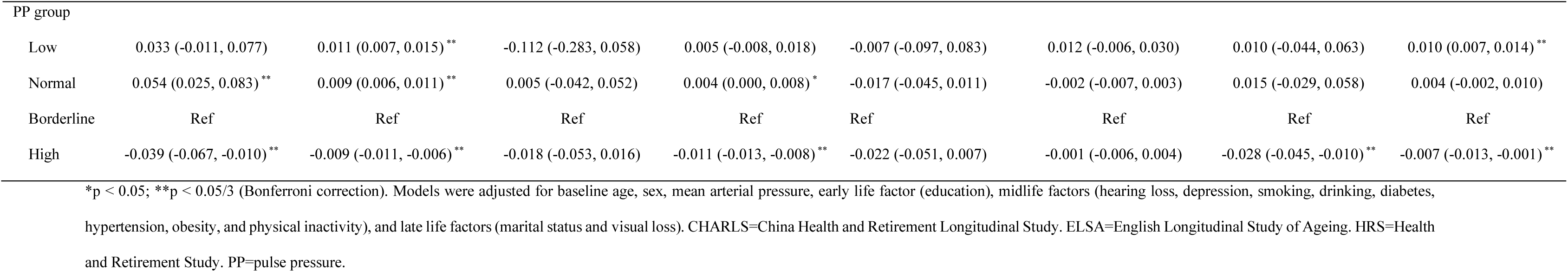
Associations of PP thresholds with cognition.

|  | HRS |  | ELSA |  | CHARLS |  | Pool |  |
| --- | --- | --- | --- | --- | --- | --- | --- | --- |
|  | Main Effect | Time Interaction Effect | Main Effect | Time Interaction Effect | Main Effect | Time Interaction Effect | Main Effect | Time Interaction Effect |
| **Memory** |  |  |  |  |  |  |  |  |
| PP Group |  |  |  |  |  |  |  |  |
| Low | -0.078 (-0.118, -0.039) ** | 0.002 (-0.003, 0.007) | -0.198 (-0.357, -0.039) | 0.005 (-0.008, 0.018) | 0.032 (-0.057, 0.120) | -0.003 (-0.020, 0.013) | -0.068 (-0.181, 0.045) | 0.002 (-0.002, 0.007) |
| Normal | Ref | Ref | Ref | Ref | Ref | Ref | Ref | Ref |
| Borderline | -0.037 (-0.065, -0.009) ** | -0.016 (-0.019, -0.012) ** | 0.008 (-0.035, 0.052) | -0.003 (-0.006, 0.001) | -0.006 (-0.034, 0.022) | -0.013 (-0.018, -0.008) ** | -0.015 (-0.041, 0.011) | -0.010 (-0.018, -0.003) ** |
| High | -0.094 (-0.124, -0.064) ** | -0.028 (-0.032, -0.025) ** | -0.013 (-0.057, 0.031) | -0.015 (-0.019, -0.012) ** | -0.072 (-0.103, -0.040) ** | -0.034 (-0.038, -0.029) ** | -0.062 (-0.107, -0.016) ** | -0.026 (-0.036, -0.015) ** |
| PP Group |  |  |  |  |  |  |  |  |
| Low | -0.041 (-0.083, 0.001) | 0.018 (0.013, 0.023) ** | -0.206 (-0.364, -0.049) ** | 0.007 (-0.006, 0.020) | 0.038 (-0.052, 0.127) | 0.010 (-0.007, 0.027) | -0.054 (-0.174, 0.066) | 0.014 (0.006, 0.022) ** |
| Normal | 0.037 (0.009, 0.065) ** | 0.016 (0.012, 0.019) ** | -0.008 (-0.052, 0.035) | 0.003 (-0.001, 0.006) | 0.006 (-0.022, 0.034) | 0.013 (0.008, 0.018) ** | 0.015 (-0.011, 0.041) | 0.010 (0.003, 0.018) ** |
| Borderline | Ref | Ref | Ref | Ref | Ref | Ref | Ref | Ref |
| High | -0.057 (-0.084, -0.030) * | -0.013 (-0.016, -0.010) ** | -0.021 (-0.053, 0.011) | -0.013 (-0.015, -0.010) | -0.066 (-0.095, -0.037) ** | -0.020 (-0.025, -0.016) ** | -0.049 (-0.074, -0.023) ** | -0.015 (-0.020, -0.010) ** |
| **Orientation** |  |  |  |  |  |  |  |  |
| PP group |  |  |  |  |  |  |  |  |
| Low | -0.059 (-0.108, -0.010) * | 0.001 (-0.008, 0.010) | -0.121 (-0.235, -0.007) * | 0.004 (-0.011, 0.020) | 0.028 (-0.067, 0.123) | -0.006 (-0.022, 0.011) | -0.048 (-0.118, 0.021) | 0.001 (-0.006, 0.008) |
| Normal | Ref | Ref | Ref | Ref | Ref | Ref | Ref | Ref |
| Borderline | 0.010 (-0.020, 0.040) | -0.013 (-0.018, -0.007) ** | 0.008 (-0.024, 0.041) | 0.001 (-0.003, 0.006) | 0.021 (-0.009, 0.051) | 0.001 (-0.004, 0.006) | 0.013 (-0.005, 0.031) | -0.003 (-0.012, 0.005) |
| High | -0.009 (-0.040, 0.021) | -0.022 (-0.027, -0.017) ** | 0.010 (-0.022, 0.042) | -0.006 (-0.010, -0.002) ** | -0.037 (-0.071, -0.003) * | -0.008 (-0.012, -0.003) ** | -0.011 (-0.037, 0.014) | -0.012 (-0.022, -0.002) ** |
| PP group |  |  |  |  |  |  |  |  |
| Low | -0.068 (-0.118, -0.019) ** | 0.014 (0.005, 0.022) ** | -0.129 (-0.242, -0.016) * | 0.003 (-0.012, 0.018) | 0.007 (-0.089, 0.103) | -0.006 (-0.023, 0.011) | -0.061 (-0.118, -0.004) | 0.005 (-0.006, 0.017) |
| Normal | -0.010 (-0.040, 0.020) | 0.013 (0.007, 0.018) ** | -0.008 (-0.041, 0.024) | -0.001 (-0.006, 0.003) | -0.021 (-0.051, 0.009) | -0.001 (-0.006, 0.004) | -0.013 (-0.031, 0.005) | 0.003 (-0.005, 0.012) |
| Borderline | Ref | Ref | Ref | Ref | Ref | Ref | Ref | Ref |
| High | -0.019 (-0.044, 0.007) | -0.010 (-0.014, -0.005) ** | 0.002 (-0.021, 0.025) | -0.008 (-0.010, -0.005) ** | -0.058 (-0.089, -0.027) ** | -0.008 (-0.013, -0.003) ** | -0.024 (-0.057, 0.010) | -0.008 (-0.010, -0.006) ** |
| **Executive Function** |  |  |  |  |  |  |  |  |
| PP group |  |  |  |  |  |  |  |  |
| Low | -0.020 (-0.062, 0.021) | 0.002 (-0.002, 0.006) | -0.117 (-0.289, 0.054) | 0.000 (-0.013, 0.014) | 0.010 (-0.080, 0.100) | 0.014 (-0.003, 0.032) | -0.020 (-0.057, 0.017) | 0.003 (-0.001, 0.007) |
| Normal | Ref | Ref | Ref | Ref | Ref | Ref | Ref | Ref |
| Borderline | -0.054 (-0.083, -0.025) ** | -0.009 (-0.011, -0.006) ** | -0.005 (-0.052, 0.042) | -0.004 (-0.008, -0.000) * | 0.017 (-0.011, 0.045) | 0.002 (-0.003, 0.007) | -0.015 (-0.058, 0.029) | -0.004 (-0.010, 0.002) |
| High | -0.093 (-0.124, -0.061) ** | -0.017 (-0.020, -0.015) ** | -0.024 (-0.071, 0.024) | -0.015 (-0.019, -0.011) ** | -0.005 (-0.037, 0.027) | 0.002 (-0.003, 0.007) | -0.041 (-0.095, 0.013) | -0.010 (-0.022, 0.001) |

| PP group |  |  |  |  |  |  |  |  |
| --- | --- | --- | --- | --- | --- | --- | --- | --- |
| Low | 0.033 (-0.011, 0.077) | 0.011 (0.007, 0.015) ** | -0.112 (-0.283, 0.058) | 0.005 (-0.008, 0.018) | -0.007 (-0.097, 0.083) | 0.012 (-0.006, 0.030) | 0.010 (-0.044, 0.063) | 0.010 (0.007, 0.014) ** |
| Normal | 0.054 (0.025, 0.083) ** | 0.009 (0.006, 0.011) ** | 0.005 (-0.042, 0.052) | 0.004 (0.000, 0.008) * | -0.017 (-0.045, 0.011) | -0.002 (-0.007, 0.003) | 0.015 (-0.029, 0.058) | 0.004 (-0.002, 0.010) |
| Borderline | Ref | Ref | Ref | Ref | Ref | Ref | Ref | Ref |
| High | -0.039 (-0.067, -0.010) ** | -0.009 (-0.011, -0.006) ** | -0.018 (-0.053, 0.016) | -0.011 (-0.013, -0.008) ** | -0.022 (-0.051, 0.007) | -0.001 (-0.006, 0.004) | -0.028 (-0.045, -0.010) ** | -0.007 (-0.013, -0.001) ** |

\*p < 0.05; \*\*p < 0.05/3 (Bonferroni correction). Models were adjusted for baseline age, sex, mean arterial pressure, early life factor (education), midlife factors (hearing loss, depression, smoking, drinking, diabetes, hypertension, obesity, and physical inactivity), and late life factors (marital status and visual loss). CHARLS=China Health and Retirement Longitudinal Study. ELSA=English Longitudinal Study of Ageing. HRS=Health and Retirement Study. PP=pulse pressure.

High PP was also associated with a greater orientation decline across three studies (pooled β = −0.012 SD/year; 95% CI: −0.022, −0.002), and with greater executive function decline in HRS (β = −0.017 SD/year; 95% CI: −0.020, −0.015) and in ELSA (β = −0.015 SD/year; 95% CI: −0.019, −0.011).

### 3.4 Associations of PP with cognition stratified by SBP, race and age

The results remained robust even among individuals with Normal SBP, with higher PP associated with poorer memory performance (pooled β = −0.071 SD; 95% CI: −0.105 to −0.037) and greater memory decline (pooled β = −0.021 SD/year; 95% CI: −0.032 to −0.011). In the High SBP group, PP remained significantly associated with greater decline in the pool analysis along with the HRS and CHARLS cohorts (pooled β = −0.021 SD/year; 95% CI: −0.035, −0.007). Stratified by race, age, and sex, higher PP was significantly associated with greater memory decline across White, Black, and Asian populations, midlife (<60 years) and older (≥60 years) groups, as well as in both male and female (Table 3).

**Table 3:**
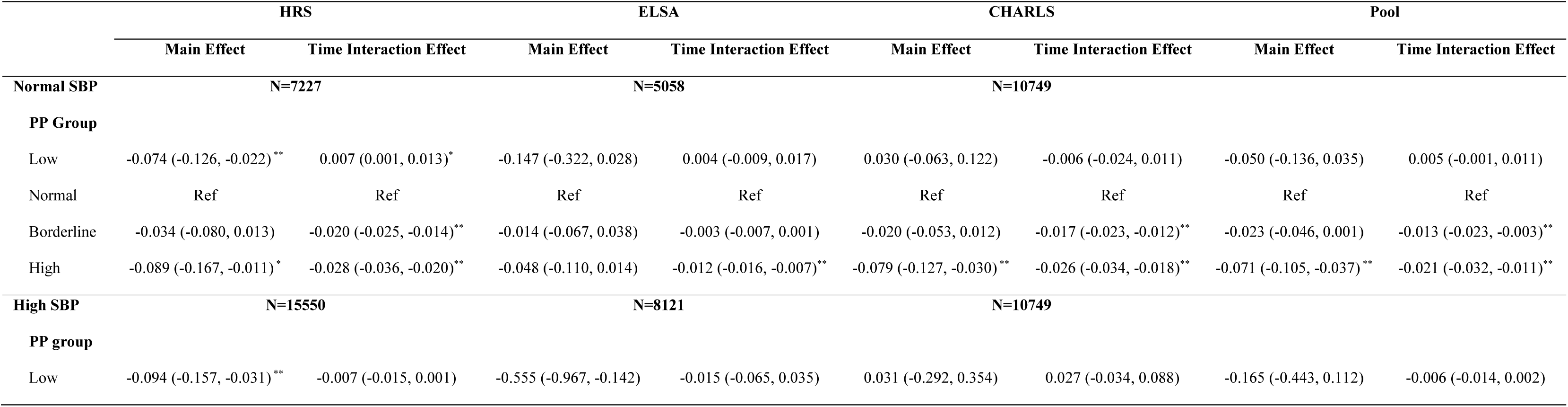
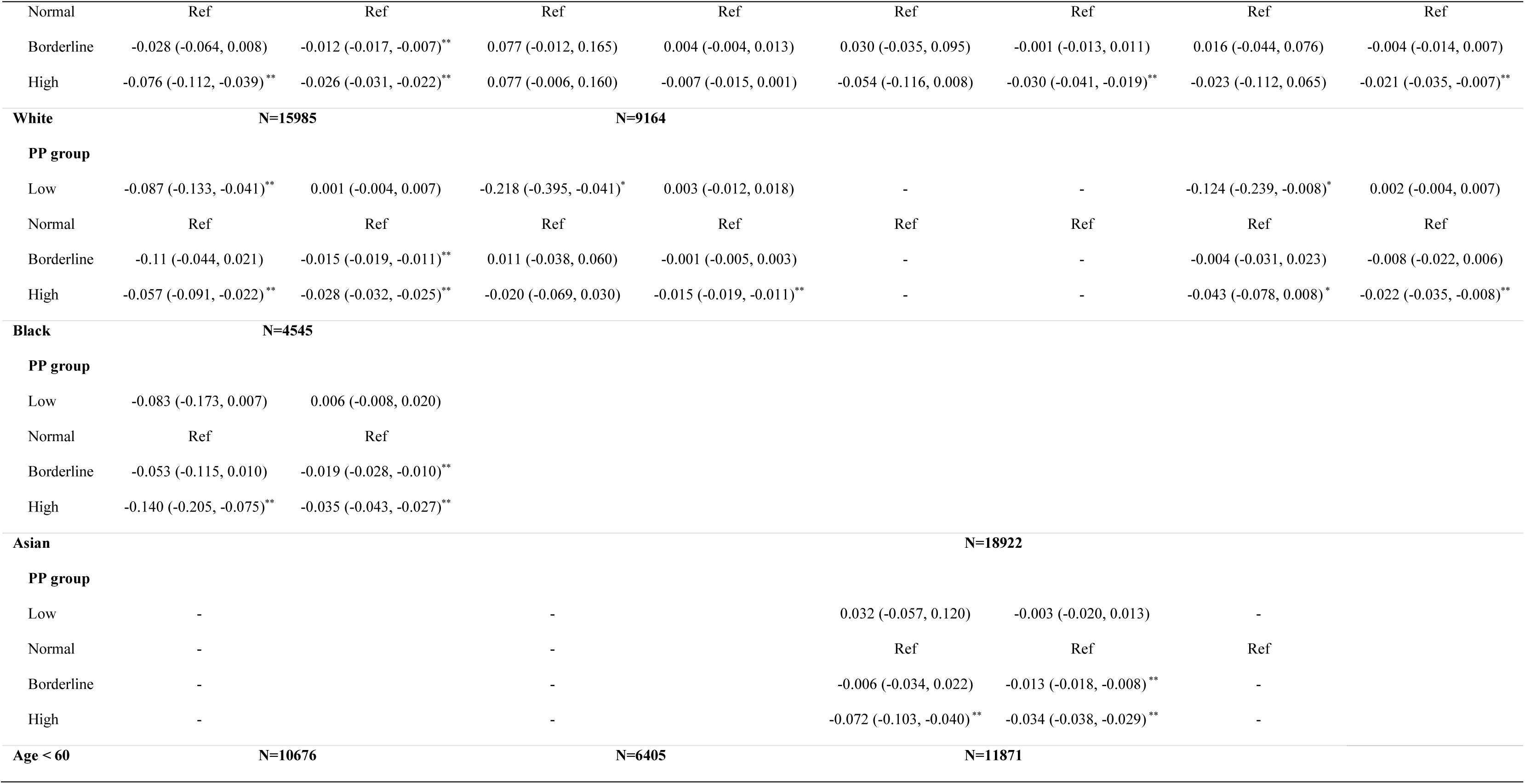
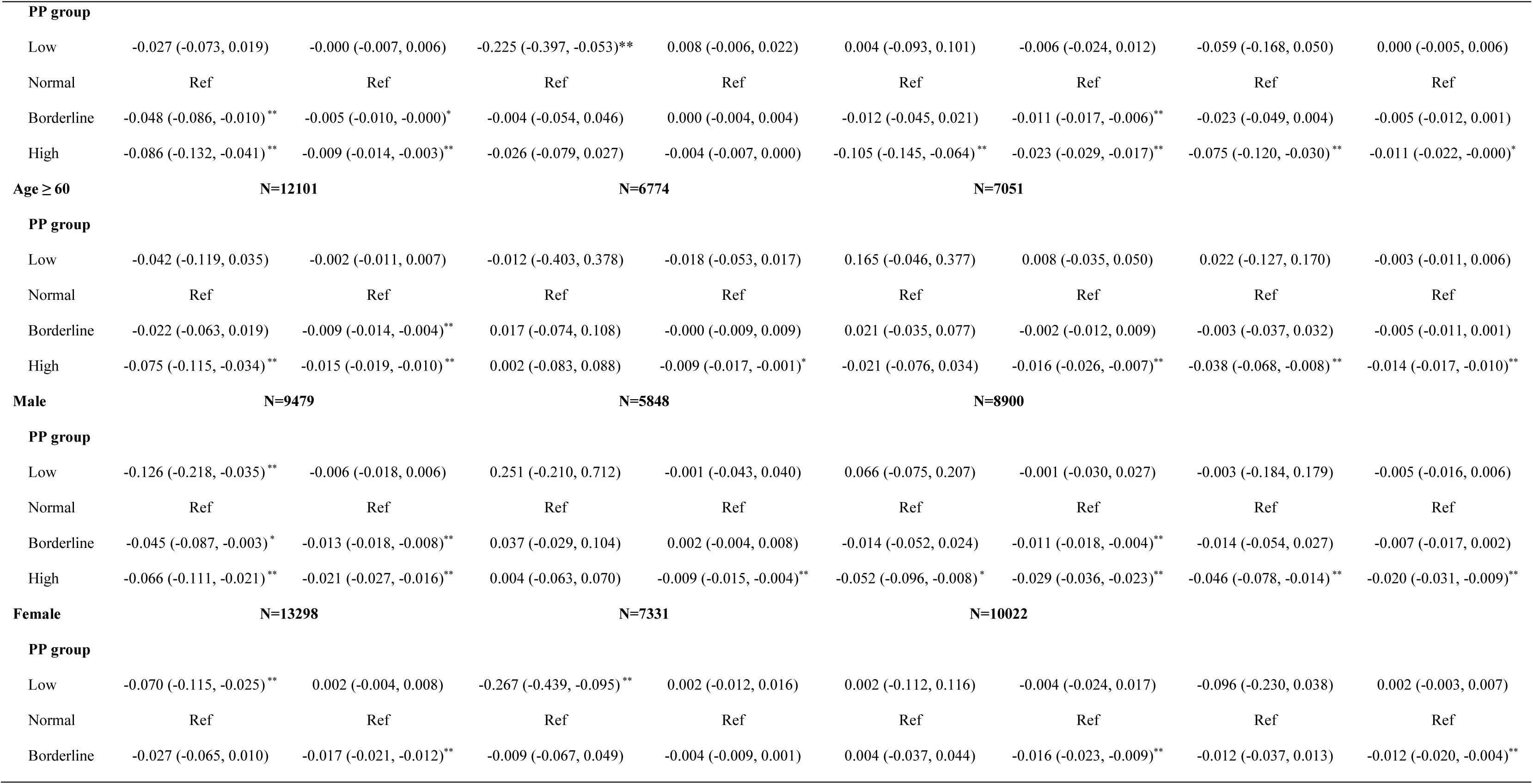
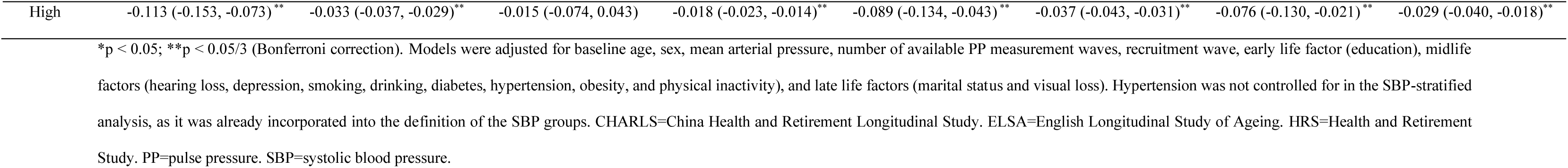
Associations of PP thresholds with Memory stratified by SBP, Race, Age, and Sex.

### 3.5 Sensitivity Analysis

The nonlinear association between PP and memory remained robust after accounting for the nonlinear effect of baseline age (Figure S4). Participants whose PP exceeded the Borderline (50 mmHg) or High (60 mmHg) threshold in a single assessment already showed greater memory decline, while those who exceeded these thresholds across multiple assessments experienced even greater decline (Table S13). The results remained robust after additionally controlling for antihypertensive medication use (Table S14), excluding those without PP follow-up assessments (Table S15), using pattern-mixture models (Table S16), excluding those with baseline stroke and heart disease (Table S17), in the complete case analysis (Table S18), as well as in one-step IPD analysis (Table S19).

## 4 Discussion

Using a longitudinal approach in ethnically- and geographically-diverse populations, we confirmed an overall nonlinear association between higher pulse pressure and poorer memory, which was more pronounced at higher levels of pulse pressure, independent of traditional blood pressure measures (mean arterial pressure or just systolic pressure), and independent of other established modifiable dementia risk factors. A pulse pressure ≥60 mmHg was associated with worse memory performance and faster memory decline, including among normotensive individuals, with consistent findings across diverse ethnic and regional populations (UK, US and China), racial groups (White, Black and Asian), age strata (midlife and late life) and both sexes.

Our results showed that each 10 mmHg increase in pulse pressure was associated with an additional 0.014 SD increase in memory decline, an effect size comparable to those of established dementia risk factors (e.g., hypertension, diabetes, and smoking), both observed in the current study (Table S20) and reported in previous literature.^23,40–42^ Previous epidemiology studies have focused on the effect of pulse pressure on cardiovascular outcomes rather than cognition.^17,19,43^ While some studies have examined its effects on cognition,^14,16^ these have focused on a global measure of cognition in a single population. Our research shows that this association is clearest in terms of episodic memory, and generalises across populations in UK, US and China, as well as across White, Black and Asian people. Although we found similar patterns, whether the effect of PP on memory differs in magnitude across ethnic groups remains unclear and warrants further investigation. In contrast, we did not find any association with a cognitive measure of “orientation” (in time), and any association that might exist with executive function was not found in all cohorts (there was a basic association with decline in executive function in the US and UK cohorts, consistent with prior reports on UK cohorts,^4,12^ but this association did not get stronger with higher levels of pulse pressure, and was not found in the Chinese cohort). The selective, or at least, stronger effect of pulse pressure on episodic memory may partly reflect the structural vulnerability of hippocampal vasculature.^44,45^ This vascular susceptibility, combined with the sensitivity of hippocampus to hypoxia^46^, may make episodic memory particularly susceptible to age-related arterial stiffening and raised pulse pressure.

We evaluated non-linear associations and validated the high pulse pressure threshold for interpretability. Across all cohorts, memory slopes were negative and significant for pulse pressure ≥60 mmHg, while values below this threshold crossed the zero interval, indicating no robust association at normal-to-lower pulse pressures. Orientation and executive function showed inconsistent non-linear effects across cohorts, consistent with null pooled results.^4,121213,47^ Overall, memory emerged as the most robust and generalisable cognitive domain for capturing effects of elevated pulse pressure, supporting its use as the primary outcome for evaluating clinically interpretable thresholds, whereas executive function appeared to reflect more complex, non-linear vascular effects.

A key contribution of the present study was the confirmation of clinically interpretable thresholds. Individuals with pulse pressure ≥60 mmHg showed significant memory impairment and greater memory decline across populations and even among nominally normotensive individuals. In addition, individuals with pulse pressure ≥50 mmHg (the borderline group) already demonstrated evidence of memory decline, consistent with the non-linear association between pulse pressure and memory, and suggesting that cognitive deterioration begins at relatively modest elevations in pulse pressure. The results are similar with previous study that reported non-linear associations between pulse pressure with cardiovascular outcomes in worldwide trials, and with cognition in Chinese populations.^43,48^ The present study expands the clinical significance of pulse pressure thresholds by demonstrating their relevance not only for cardiovascular outcomes, but also for domain-specific cognitive decline.^18^

There are some suggestions about the biological and vascular mechanisms by which pulse pressure affects cognition.^12^ Pulse pressure, a surrogate measure of arterial stiffness, leads to increased transmission of pulsatile hemodynamic forces to the microvasculature, contributing to structural brain aging and subsequent cognitive decline.^49,50^ Previous research has elucidated that white matter microstructure impairment mediates the effect of pulse pressure onto cognition.^12^ For example, there was a curvilinear association between pulse pressure and white matter microstructure injuries, with the curve relatively flat at normal pulse pressures and steepening nearly exponentially at higher pressures.^12^ The threshold for an effect of pulse pressure to be manifest may arise after a failure of compensatory mechanisms. Small fluctuations within normal pulse pressure can be buffered by vessel elasticity and the Windkessel effect, whereas pressures exceeding the compensatory limits penetrate the microcirculation, leading to cerebral damage, disruption of white matter integrity, and impairment of cerebral blood flow autoregulation.^12,13^

### Clinical and Public Health Implications

The present study carries important clinical and public health implications, supporting the use of pulse pressure thresholds as a clinically interpretable and complementary marker to systolic blood pressure for assessing the risk of memory decline across diverse populations. Pulse pressure, calculated by systolic and diastolic blood pressure, is routinely available in clinical practice and at home, and is relevant for midlife prevention strategies.^3^ The association between pulse pressure and cognition in normotensive individuals highlights a limitation of current systolic pressure risk stratification in clinical practice (e.g. >140mmHg for hypertensive treatment). Elevated pulse, even in the presence of normal SBP, may identify individuals at risk of cognitive decline and dementia who would otherwise remain undetected.^43^ Targeting pulse pressure may therefore represent a therapeutic strategy beyond conventional blood pressure management, potentially facilitating early dementia prevention.

### Equity and Generalisability

The present study estimated vascular-cognitive relationships across multiple ethnoregional cohorts (UK, US and China), racial groups (White, Black and Asian), age strata (midlife and late life), and sexes, enhancing the equity and generalizability of the findings. While pulse pressure consistently contributes memory across populations, its association with executive function decline was observed mainly in US and UK populations, aligning with prior research demonstrating similar findings in non-Asian populations.^4,12,49^ Therefore, pulse pressure may serve as a practical target for dementia prevention worldwide, with consideration of racial and ethnic differences. The effect of pulse pressure on executive function may be specific to non-Asian populations, and the mechanisms underlying this discrepancy require further exploration.

### Strengths and Limitations

The study has several strengths. To our knowledge, this is the first study to characterise pulse pressure thresholds for cognitive outcomes, providing insight for identifying an earlier management window for cognitive decline and dementia. Secondly, the study harmonized large and diverse longitudinal cohorts from different regions and ethnic groups, thus demonstrating the equity and generalizability of findings. Thirdly, cognitive measures across diverse cognitive domains were statistically harmonised, allowing for an in-depth, domain-specific investigation. Fourthly, non-linear and threshold-based analyses, grounded in vascular aging theory and established guidelines, enabled the identification of clinically interpretable pulse pressure cutoffs, thereby enhancing clinical and public health relevance. Fifthly, the present study comprehensively adjusted for traditional blood pressure measures and other established dementia risk factors, highlighting the independent and incremental prognostic value of pulse pressure for cognitive outcomes.

This study also has several limitations. Firstly, causal relationships between pulse pressure and cognition cannot be fully elucidated from observation data. Secondly, pulse pressure derived from brachial blood pressure is not a direct measure of arterial stiffness and may be mixed with other mechanisms. Ideally, future studies should adopt a multimodal approach, such as integration with vascular and neuroimaging data, to better elucidate these pathways. Thirdly, memory and orientation tasks were harmonised across cohorts, while executive function in ELSA relied on study-specific task, which despite statistical harmonisation, may have contributed to additional measurement variability. However, for executive function, HRS and ELSA showed consistent results despite using different tasks, whereas HRS and the China cohort showed inverse results despite using the same tasks, suggesting that task differences are unlikely to explain the observed variability. In addition, the limited range of orientation tasks may reduce the sensitivity to detect differences. Fourthly, residual confounding is possible, and spontaneous variability in pulse pressure measures, including single-wave measurements and white-coat effects, may affect threshold estimation, although these concerns were mitigated through sensitivity analyses. Pulse pressure, as a surrogate of arterial stiffness, is likely more stable than systolic pulse pressure, which may explain its significant effects on cognition and supporting its clinical relevance. Lastly, ethnicity-specific thresholds across different time ranges warrant further validation.

## Conclusions

This study shows that pulse pressure above 60 mmHg identified individuals at elevated cognitive risk, over and above systolic and diastolic blood pressures, and even among normotensive individuals. Early prevention of cognitive decline could be guided by monitoring for and treating pulse pressure above 50 mmHg. These results support consideration of pulse pressure thresholds as a clinically interpretable and complementary marker for dementia risk assessment across diverse populations.

## Contributors

HRZ and KAT accessed and verified the data. KAT, XX, and HRZ designed the study. HRZ, SHC, and KAT analysed the data. HRZ, SHC, HXW, and KAT interpreted the results and drafted the manuscript and prepared the tables and figures. The manuscript and figures were critically reviewed and revised by all authors. KAT and XX are joint senior authors.

## Acknowledgements

K.A.T. was supported by a Fellowship award from the Alzheimer’s Society, UK (Grant number 602). J.B.R was supported by the NIHR Cambridge Biomedical Research Centre (NIHR203312: BRC-1215-20014), NIHR funding to the NIHR BioResource (RG94028 & RG85445), Wellcome Trust (220258), Medical Research Council (SUAG/051G101400; SUAG/010 RG91365; MC_UU_00030/14 and MR/T033371/1), the Holt Fellowship and by the Addenbrookes Charitable Trust. We thank NIHR BioResource volunteers for their participation, and gratefully acknowledge NIHR BioResource centres, NHS Trusts and staff for their contribution. The views expressed are those of the author(s) and not necessarily those of the NHS or the NIHR or the Department of Health and Social Care. XX was funded by Natural Science Foundation of China (NSFC/72274170, NSFC/82201733). The funders had no role in the design and conduct of the study; collection, management, analysis, and interpretation of the data; preparation, review, or approval of the manuscript; and decision to submit the manuscript for publication. The authors appreciate all participants for their involvement and efforts made by the original data creators.

## Data sharing

Original survey datasets from the HRS, ELSA, and CHARLS are freely available to all bona fide researchers. Access to data can be obtained by visiting their web sites. The data can also be obtained upon request to the corresponding author.

