## Supplementary Material for "Pulse pressure thresholds associated with cognitive impairment across diverse regional and ethnic populations"

**Supplementary Materials**

**Supplementary Methods**

**Table S1: Definition of Pulse Pressure Group**

**Table S2: Definition of Cognitive Assessments**

**Table S3: Definition of modifiable factors of dementia across studies**

**Table S4: Number and percentage of missing data**

**Table S5: Baseline characteristics of participants in HRS**

**Table S6: Baseline characteristics of participants in ELSA**

**Table S7: Baseline characteristics of participants in CHARLS**

**Table S8: Model fit summary**

**Table S9: Model fit and comparison for Restricted Cubic Spline Models with 3–5 Knots**

**Table S10: Baseline characteristics of participants in different PP groups in HRS**

**Table S11: Baseline characteristics of participants in different PP groups in ELSA**

**Table S12: Baseline characteristics of participants in different PP groups in CHARLS**

**Table S13: Associations of PP thresholds and frequency of PP wave exceedance with cognition**

**Table S14: Associations of PP thresholds with cognition considering antihypertensive medication use**

**Table S15: Associations of PP thresholds with cognition among participants with PP follow-up assessments**

**Table S16: Associations of PP thresholds with cognition among participants based on pattern-mixture models**

**Table S17: Associations of PP thresholds with cognition among participants without baseline stroke and heart disease**

**Table S18: Associations of PP thresholds with cognition in complete case analysis**

**Table S19: Associations of PP thresholds with cognition in one-step IPD analysis**

**Table S20: Associations of baseline hypertension, diabetes, and smoking with memory**

**Figure S1: Nonlinear associations of PP with memory (A-C), orientation (D-F), and execution (G-I) in HRS (A, D, G), ELSA (B, E, H), and CHARLS (C, F, I)**

**Figure S2: Nonlinear associations of PP baseline and change with memory (A-C), orientation (D-F), and execution (G-I) in HRS (A, D, G), ELSA (B, E, H), and CHARLS (C, F, I)**

**Figure S3: Associations of PP with Memory stratified by PP threshold groups**

**Figure S4: Associations of PP baseline and change with memory**

**Supplementary Methods:**

We performed a series of regression models from simple to complex, only retaining variables that benefited model fit. Model fit was investigated using the Akaike Information Criterion (AIC), Bayesian Information Criterion (BIC), and marginal variance explained (R^2^), with better fit indicated by lower AIC and BIC values and higher marginal R^2^. Statistical significance was defined as *P*<0.05.

In total, five linear mixed-effect regression models tested two sets of nested hypotheses after establishing the baseline model. The baseline model was fitted to establish the effect of age on cognition, serving as reference against which the independent association between PP and cognition was evaluated:

*Model 1: Cognition ~ Age + (1|Participant)*

The first set of models (Models 2A-B) sought to assess the between-person and within-person effects of PP on cognition, assuming these were independent of age. Model 2A examined the linear effects of PP specified as follows:

*Model 2A:* *Cognition ~ PP + Age + (1|Participant)*

Model 2B assessed the presence of nonlinear effects of PP, given previous evidence suggesting potential threshold effects. To this end, we applied restricted cubic splines to characterise the functional form of this relationship and inform subsequent threshold analyses (Model 2B).^33^

*Model 2B:* *Cognition ~ Spline (PP) + Age + (1|Participant)*

Models 2A and 2B were compared and if splines improved fit, they were taken into Models 3A-B. Results were presented for Models 2C. Covariates included sex, MAP, early, midlife, and late-life of modifiable factors.

*Model 2C: Cognition ~ Spline (PP) + Age + Covariates + (1|Participant)*

Models 3A-B tested the nonlinear association between PP and cognition in longitudinal data, allowing for distinct between-participant and within-participant effects. To address this, PP was decomposed into a between-person component (baseline PP) and a within-person component (change in PP over time), which were allowed to interact. This tests the hypothesis that within-person association between PP change and cognition may vary as a function of baseline PP (See model 3A).

*Model 3A: Cognition ~ Spline (PP baseline) * Spline (PP change) + Age + (1|Participant)*

This specification allows for a fully flexible, non-linear interaction between the between-person (baseline PP) and within-person (PP change) components.^34^ Baseline PP captures stable between-person differences in pulse pressure and is expected to span the full non-linear range of the hypothesized inverted-U or J-shaped association. However, over a relatively short follow-up period (2–4-years), within-person changes in PP are typically modest at the population level^35^. As such, the within-person effect of PP is unlikely to be well captured by a highly flexible non-linear specification. To enhance interpretability and parsimony, we therefore fitted a simplified model in which only the between-person component was modelled non-linearly, while the within-person component was specified as a linear effect (Model 3B):

*Model 3B: Cognition ~ Spline (PP baseline) * PP change + Age + (1|Participant)*

In this specification, the interaction term allows the effect of PP change on cognition to vary across levels of baseline PP, thereby capturing potential non-linearity while maintaining interpretability. The optimal number of knots for the restricted cubic splines was evaluated by fitting models with three to five knots.

Finally, to accommodate the covariates, results were also compared between Model 2C and 3C:

*Model 3C: Cognition ~ Spline (PP baseline) * PP change + Age + Covariates + (1|Participant)*

**Table S1: Definition of Pulse Pressure Group**

| **Group** | **Definition** | **Abbreviation** |
| --- | --- | --- |
| Low | At least one wave has a PP value below 30 mmHg, and the maximum PP value does not exceed 50 mmHg. | <30 |
| Normal | All waves have PP values ranging from 30 mmHg to < 50 mmHg. | 30-50 |
| Borderline | At least one wave has a PP value of 50 mmHg or higher, and all waves have PP values below 60 mmHg. | <60 |
| High | At least one wave has a value of 60 mmHg or higher. | ≥60 |
| PP=Pulse Pressure. | | |

**Table S2: Definition of Cognitive Assessments**

|  |  | **CHARLS** | **HRS** | **ELSA** |
| --- | --- | --- | --- | --- |
| Memory | Assessment | Immediate and delayed word recall:  The participants were read a set of 10 words and were asked to recall the words as many as they can, in any order (Immediate recall). After conducting some other tests, the participants were again asked to recall the words they remembered (Delayed recall). | | |
|  | Score range | 0-10 (average immediate and delayed recall） | | |
| Orientation | Assessment | Today’s date and the current season:  (day, day of the week, month, year and season) | Today’s date:  (day, day of week, month and year) | Today’s date:  (day, day of week, month and year) |
|  | Score range | 0-5 | 0-4 | 0-4 |
| Executive | Assessment | Serial 7s subtraction:  subtract 7 from the prior number,  beginning with 100 for five trials. | Serial 7s subtraction:  subtract 7 from the prior number, beginning with 100 for five trials. | Animal-naming:  list as many animal names as the participant could in 60 seconds. |
|  | Score range | 0-5 | 0-5 | Total count of words:  (After excluding repeated and non-animal words, no upper limits) |
| CHARLS=China Health and Retirement Longitudinal Study. HRS=**Health and Retirement Study.** ELSA=**English Longitudinal Study of Ageing.** | | | | |

**Table S3: Definition of modifiable factors of dementia across studies**

| **Stage** | **Modifiable risk factors** | **HRS** | **ELSA** | **CHARLS** |
| --- | --- | --- | --- | --- |
| Early Life | Less education | Years of education: | Highest level of education | Highest level of education |
|  |  | None and primary: <7  Secondary and above: ≥7 | None and primary: No qualification, NVQ1 / CSE other grade equivalent  Secondary and above: NVQ2/GCE O Level equiv, NVQ3/GCE A Level equiv, NVQ4/NVQ5/Degree or equiv, Higher education below degree, Foreign/other | None and primary: below elementary  school  Secondary and above: middle, high school, and vocational school, above college/associate degree |
| Midlife | Hearing loss | Self-report of hearing: Poor/fair | Self-report of hearing: Poor/fair | Self-report of hearing loss |
|  | High LDL cholesterol | - | | |
|  | Depression | CESD-8 score ≥ 4 | CESD-8 score ≥ 4 | CESD-10 score ≥ 12 |
|  | Traumatic brain injury | - | | |
|  | Physical inactivity | No moderate or vigorous activity on a weekly basis | No moderate or vigorous activity on a weekly basis | Physical disabilities |
|  | Diabetes | Self-report diabetes | | |
|  | Smoking | Ever smoke | | |
|  | Hypertension | Self-report high BP | | |
|  | Obesity | BMI≥28.0 | | |
|  | Excessive alcohol | Ever drink any alcohol | Ever taken an alcoholic drink in the past 12 months | Ever drink any alcoholic beverages last year |
| Late life | Social isolation | Marital status: unmarried (including separated, divorced, widowed, or never married) | | |
|  | Air pollution | - | | |
|  | Visual loss | Self-report of eyesight: poor/fair | Self-report of eyesight: poor or fair | Self-report of visual problem |
| CHARLS=China Health and Retirement Longitudinal Study. HRS=**Health and Retirement Study.** ELSA=**English Longitudinal Study of Ageing.** NVQ1=National Vocational Qualification Level. CSE=Certificate of Secondary Education. NVQ2=National Vocational Qualification Level 2. GCE O=General Certificate of Education–Ordinary Level. NVQ3=National Vocational Qualification Level 3. GCE A=General Certificate of Education–Advanced Level. NVQ4=National Vocational Qualification Level 4. NVQ5= National Vocational Qualification Level 5. CESD-8=Center for Epidemiologic Studies Depression Scale–8 item version. CESD-10=Center for Epidemiologic Studies Depression Scale–10 item version. BP=blood pressure. BMI=body mass index. Marital status was used as a proxy for social isolation due to the lack of comparable measures across studies. | | | | |

**Table S4: Number and percentage of missing data**

| **Characteristic** | **HRS (N=22777)** | **ELSA (N=13179)** | **CHARLS (N=18922)** |
| --- | --- | --- | --- |
| Age | 0 (0%) | 0 (0%) | 0 (0%) |
| Sex | 0 (0%) | 0 (0%) | 0 (0%) |
| Education | 91 (0.4%) | 72 (0.5%) | 3273 (17.3%) |
| Marital status | 9 (0.0%) | 250 (1.9%) | 1 (0.0%) |
| Smoking | 104 (0.5%) | 2542 (19.3%) | 1074 (5.7%) |
| Drinking | 4 (0.0%) | 609 (4.6%) | 4 (0.0%) |
| Hypertension | 261 (1.1%) | 881 (6.7%) | 68 (0.4%) |
| Diabetes | 158 (0.7%) | 4435 (33.7%) | 118 (0.6%) |
| Obesity | 1339 (5.9%) | 1248 (9.5%) | 209 (1.1%) |
| Hearing loss | 7 (0.0%) | 2 (0.0%) | 166 (0.9%) |
| Visual loss | 82 (0.4%) | 0 (0%) | 95 (0.5%) |
| Depression | 2 (0.0%) | 9 (0.1%) | 859 (4.5%) |
| Physical inactivity | 7 (0.0%) | 3 (0.0%) | 111 (0.6%) |
| CHARLS=China Health and Retirement Longitudinal Study. HRS=**Health and Retirement Study.** ELSA= **English Longitudinal Study of Ageing.** | | | |

**Table S5: Baseline characteristics of participants in HRS**

| **Characteristic** | **Baseline Wave** | | | | | | | **Total** |
| --- | --- | --- | --- | --- | --- | --- | --- | --- |
|  | **Wave 0** | **Wave 1** | **Wave 2** | **Wave 3** | **Wave 4** | **Wave 5** | **Wave 6** |  |
| N | 6828 | 5823 | 3174 | 3126 | 624 | 1791 | 1411 | 22777 |
| Recruit time, year | 2006-2007 | 2008-2009 | 2010-2011 | 2012-2013 | 2014-2015 | 2016-2017 | 2018-2019 | - |
| Age, mean (SD), year | 67.7 (10.4) | 68.9 (9.87) | 57.6 (8.62) | 58.7 (8.20) | 63.1 (9.82) | 55.4 (6.79) | 56.2 (5.87) | 63.6 (10.7) |
| Sex, male, % | 2795 (40.9%) | 2303 (39.6%) | 1347 (42.4%) | 1362 (43.6%) | 288 (46.2%) | 788 (44.0%) | 596 (42.2%) | 9479 (41.6%) |
| Ethnicity, white, % | 5605 (82.1%) | 4755 (81.7%) | 1775 (55.9%) | 1785 (57.1%) | 394 (63.1%) | 936 (52.3%) | 735 (52.1%) | 15985 (70.2%) |
| SBP, mean (SD), mmHg | 131 (20.6) | 131 (20.2) | 128 (20.8) | 127 (20.2) | 128 (21.6) | 126 (19.5) | 124 (18.7) | 129 (20.4) |
| PP, mean (SD), mmHg | 51.2 (14.6) | 51.7 (14.5) | 46.2 (13.5) | 46.1 (13.1) | 48.5 (14.3) | 44.2 (13.4) | 43.9 (12.7) | 48.9 (14.3) |
| MAP, mean (SD), mmHg | 96.4 (13.6) | 96.4 (13.2) | 97.7 (14.3) | 96.6 (13.9) | 95.9 (14.5) | 96.3 (13.4) | 95.1 (12.9) | 96.5 (13.6) |
| Modifiable factors (%) | | | | | | | | |
| Education | | | | | | | | |
| None and primary | 292 (4.3%) | 269 (4.6%) | 225 (7.1%) | 199 (6.4%) | 42 (6.7%) | 91 (5.1%) | 81 (5.7%) | 1199 (5.3%) |
| Secondly and above | 6524 (95.5%) | 5547 (95.3%) | 2925 (92.2%) | 2889 (92.4%) | 575 (92.1%) | 1697 (94.8%) | 1330 (94.3%) | 21487 (94.3%) |
| Marital status, unmarried | 2092 (30.6%) | 1922 (33.0%) | 1025 (32.3%) | 1028 (32.9%) | 156 (25.0%) | 616 (34.4%) | 471 (33.4%) | 7310 (32.1%) |
| Smoking | 3801 (55.7%) | 3284 (56.4%) | 1788 (56.3%) | 1783 (57.0%) | 348 (55.8%) | 954 (53.3%) | 698 (49.5%) | 12656 (55.6%) |
| Drinking | 3653 (53.5%) | 3070 (52.7%) | 2077 (65.4%) | 1943 (62.2%) | 348 (55.8%) | 1265 (70.6%) | 920 (65.2%) | 13276 (58.3%) |
| Hypertension | 3769 (55.2%) | 3433 (59.0%) | 1620 (51.0%) | 1619 (51.8%) | 378 (60.6%) | 854 (47.7%) | 690 (48.9%) | 12363 (54.3%) |
| Diabetes | 1294 (19.0%) | 1186 (20.4%) | 642 (20.2%) | 662 (21.2%) | 157 (25.2%) | 388 (21.7%) | 314 (22.3%) | 4643 (20.4%) |
| Obesity | 3453 (50.6%) | 3020 (51.9%) | 1802 (56.8%) | 1829 (58.5%) | 361 (57.9%) | 1045 (58.3%) | 859 (60.9%) | 12369 (54.3%) |
| Hearing loss | 1454 (21.3%) | 1158 (19.9%) | 489 (15.4%) | 487 (15.6%) | 123 (19.7%) | 265 (14.8%) | 195 (13.8%) | 4171 (18.3%) |
| Visual loss | 1342 (19.7%) | 1171 (20.1%) | 850 (26.8%) | 824 (26.4%) | 178 (28.5%) | 501 (28.0%) | 391 (27.7%) | 5257 (23.1%) |
| Depression | 935 (13.7%) | 711 (12.2%) | 606 (19.1%) | 549 (17.6%) | 111 (17.8%) | 335 (18.7%) | 254 (18.0%) | 3501 (15.4%) |
| Physical inactivity | 423 (6.2%) | 361 (6.2%) | 196 (6.2%) | 167 (5.3%) | 55 (8.8%) | 83 (4.6%) | 58 (4.1%) | 1343 (5.9%) |
| HRS=**Health and Retirement Study**=SD, standard deviation. SBP=systolic blood pressure. PP=pulse pressure. MAP= mean arterial pressure. | | | | | | | | |

**Table S6: Baseline characteristics of participants in ELSA**

| **Characteristic** | **Baseline Wave** | | | | | | **Total** |
| --- | --- | --- | --- | --- | --- | --- | --- |
|  | **Wave 0** | **Wave 1** | **Wave 3** | **Wave 5** | **Wave 7** | **Wave 8** |  |
| N | 7465 | 1265 | 2705 | 1013 | 11 | 720 | 13179 |
| Recruit time, year | 2002-2003 | 2004-2005 | 2008-2009 | 2012-2013 | 2016-2017 | 2018-2019 | - |
| Age, mean (SD), year | 64.1 (10.2) | 66.9 (9.28) | 60.6 (7.54) | 57.6 (7.08) | 56.9 (7.56) | 55.1 (4.34) | 62.7 (9.64) |
| Sex, male, % | 3226 (43.2%) | 581 (45.9%) | 1274 (47.1%) | 462 (45.6%) | 5 (45.5%) | 300 (41.7%) | 5848 (44.4%) |
| Ethnicity, white, % | 3708 (49.7%) | 1239 (97.9%) | 2601 (96.2%) | 957 (94.5%) | 11 (100%) | 648 (90.0%) | 9164 (69.5%) |
| SBP, mean (SD), mmHg | 140 (20.0) | 136 (19.1) | 131 (17.0) | 129 (17.3) | 130 (19.9) | 125 (16.3) | 136 (19.7) |
| PP, mean (SD), mmHg | 63.1 (15.0) | 60.6 (15.9) | 54.5 (13.1) | 51.8 (12.8) | 51.9 (14.0) | 48.7 (10.7) | 59.5 (15.2) |
| MAP, mean (SD), mmHg | 98.3 (13.5) | 95.9 (12.3) | 94.2 (11.5) | 94.0 (11.9) | 95.2 (12.7) | 92.6 (11.9) | 96.6 (13.0) |
| Modifiable factors (%) | | | | | | | |
| Education | | | | | | | |
| None and primary | 3213 (43.0%) | 493 (39.0%) | 737 (27.2%) | 210 (20.7%) | 2 (18.2%) | 74 (10.3%) | 4729 (35.9%) |
| Secondly and above | 4249 (56.9%) | 770 (60.9%) | 1966 (72.7%) | 800 (79.0%) | 9 (81.8%) | 584 (81.1%) | 8378 (63.6%) |
| Marital status, unmarried | 2283 (30.6%) | 482 (38.1%) | 982 (36.3%) | 433 (42.7%) | 3 (27.3%) | 297 (41.3%) | 4480 (34.0%) |
| Smoking | 4725 (63.3%) | 2 (0.2%) | 1023 (37.8%) | 372 (36.7%) | 0 (0%) | 234 (32.5%) | 6356 (48.2%) |
| Drinking | 6679 (89.5%) | 1021 (80.7%) | 2243 (82.9%) | 771 (76.1%) | 8 (72.7%) | 578 (80.3%) | 11300 (85.7%) |
| Hypertension | 2729 (36.6%) | 524 (41.4%) | 769 (28.4%) | 289 (28.5%) | 3 (27.3%) | 121 (16.8%) | 4435 (33.7%) |
| Diabetes | 469 (6.3%) | 112 (8.9%) | 180 (6.7%) | 72 (7.1%) | 1 (9.1%) | 47 (6.5%) | 881 (6.7%) |
| Obesity | 2932 (39.3%) | 525 (41.5%) | 1193 (44.1%) | 472 (46.6%) | 0 (0%) | 0 (0%) | 5122 (38.9%) |
| Hearing loss | 1471 (19.7%) | 262 (20.7%) | 449 (16.6%) | 171 (16.9%) | 2 (18.2%) | 81 (11.3%) | 2436 (18.5%) |
| Visual loss | 1022 (13.7%) | 184 (14.5%) | 277 (10.2%) | 113 (11.2%) | 2 (18.2%) | 76 (10.6%) | 1674 (12.7%) |
| Depression | 1120 (15.0%) | 200 (15.8%) | 379 (14.0%) | 182 (18.0%) | 1 (9.1%) | 99 (13.8%) | 1981 (15.0%) |
| Physical inactivity | 1158 (15.5%) | 194 (15.3%) | 312 (11.5%) | 142 (14.0%) | 1 (9.1%) | 74 (10.3%) | 1881 (14.3%) |
| ELSA=**English Longitudinal Study of Ageing.** SD=standard deviation. SBP, systolic blood pressure. PP=pulse pressure. MAP=mean arterial pressure. | | | | | | | |

**Table S7: Baseline characteristics of participants in CHARLS**

| **Characteristic** | **Baseline Wave** | | | **Total** |
| --- | --- | --- | --- | --- |
|  | **Wave 0** | **Wave 1** | **Wave 2** |  |
| N | 12345 | 3179 | 3398 | 18922 |
| Recruit time, year | 2011-2012 | 2013-2014 | 2015-2016 | - |
| Age, mean (SD), year | 58.8 (9.43) | 57.3 (9.87) | 52.7 (9.80) | 57.4 (9.84) |
| Sex, male, % | 5749 (46.6%) | 1516 (47.7%) | 1635 (48.1%) | 8900 (47.0%) |
| SBP, mean (SD), mmHg | 129 (21.3) | 129 (20.5) | 124 (18.3) | 128 (20.8) |
| PP, mean (SD), mmHg | 53.7 (14.8) | 52.5 (14.3) | 48.5 (12.4) | 52.6 (14.5) |
| MAP, mean (SD), mmHg | 93.3 (14.2) | 94.3 (13.9) | 91.4 (12.8) | 93.1 (13.9) |
| Modifiable factors (%) | | | | |
| Education | | | | |
| None and primary | 8362 (67.7%) | 1155 (36.3%) | 835 (24.6%) | 10352 (54.7%) |
| Secondly | 3982 (32.3%) | 683 (21.5%) | 632 (18.6%) | 5297 (28.0%) |
| Marital status, unmarried | 1417 (11.5%) | 332 (10.4%) | 226 (6.7%) | 1975 (10.4%) |
| Smoking | 4789 (38.8%) | 712 (22.4%) | 855 (25.2%) | 6356 (33.6%) |
| Drinking | 4049 (32.8%) | 1232 (38.8%) | 1361 (40.1%) | 6642 (35.1%) |
| Hypertension | 2827 (22.9%) | 662 (20.8%) | 287 (8.4%) | 3776 (20.0%) |
| Diabetes | 672 (5.4%) | 161 (5.1%) | 82 (2.4%) | 915 (4.8%) |
| Obesity | 1400 (11.3%) | 455 (14.3%) | 503 (14.8%) | 2358 (12.5%) |
| Hearing loss | 979 (7.9%) | 172 (5.4%) | 175 (5.2%) | 1326 (7.0%) |
| Visual loss | 783 (6.3%) | 145 (4.6%) | 127 (3.7%) | 1055 (5.6%) |
| Depression | 4827 (39.1%) | 972 (30.6%) | 967 (28.5%) | 6766 (35.8%) |
| Physical inactivity | 474 (3.8%) | 111 (3.5%) | 120 (3.5%) | 705 (3.7%) |
| CHARLS=China Health and Retirement Longitudinal Study. SD=standard deviation. SBP=systolic blood pressure. PP=pulse pressure. MAP=mean arterial pressure. | | | | |

**Table S8: Model fit summary**

|  | HRS | | | | | ELSA | | | | | CHARLS | | | | |
| --- | --- | --- | --- | --- | --- | --- | --- | --- | --- | --- | --- | --- | --- | --- | --- |
|  | AIC | BIC | χ^2^ | P | Marginal R^2^ | AIC | BIC | χ^2^ | P | Marginal R^2^ | AIC | BIC | χ^2^ | P | Marginal R^2^ |
| **Memory** | | | | | | | | | | | | | | | |
| Model 1 | 131749 | 131784 |  |  | 0.107 | 94352 | 94386 |  |  | 0.103 | 98118 | 98152 |  |  | 0.109 |
| Model 2A | 131684 | 131729 | 66.6 | <0.001 | 0.109 | 94291 | 94333 | 63.0 | <0.001 | 0.107 | 98109 | 98151 | 10.9 | <0.001 | 0.109 |
| Model 2B | 131662 | 131715 | 24.3 | <0.001 | 0.109 | 94292 | 94343 | 1.0 | 0.325 | 0.107 | 98102 | 98153 | 8.9 | 0.003 | 0.109 |
| Model 3B | 131510 | 131589 | 158.3 | <0.001 | 0.118 | 94080 | 94156 | 217.9 | <0.001 | 0.124 | 98089 | 98166 | 18.8 | <0.001 | 0.110 |
| Model 3A | 131434 | 131540 | 81.7 | <0.001 | 0.117 | 94023 | 94125 | 63.0 | <0.001 | 0.127 | 98068 | 98170 | 27.5 | <0.001 | 0.109 |
| Model 3C | 128452 | 128646 | 2998.9 | <0.001 | 0.202 | 91645 | 91832 | 2461.1 | <0.001 | 0.235 | 95662 | 95850 | 2425.3 | <0.001 | 0.195 |
| **Orientation** | | | | | | | | | | | | | | | |
| Model 1 | 87705 | 87738 |  |  | 0.019 | 96793 | 96827 |  |  | 0.022 | 81579 | 81612 |  |  | 0.034 |
| Model 2A | 87706 | 87747 | 1.3 | 0.254 | 0.019 | 96794 | 96837 | 0.4 | 0.519 | 0.022 | 81563 | 81605 | 17.4 | <0.001 | 0.034 |
| Model 2B | 87675 | 87725 | 32.4 | <0.001 | 0.020 | 96796 | 96847 | 0.2 | 0.672 | 0.022 | 81556 | 81606 | 8.7 | 0.003 | 0.035 |
| Model 3B | 87671 | 87746 | 10.2 | 0.017 | 0.021 | 96759 | 96835 | 43.5 | <0.001 | 0.024 | 81546 | 81621 | 16.2 | 0.001 | 0.036 |
| Model 3A | 87559 | 87659 | 117.9 | <0.001 | 0.024 | 96748 | 96850 | 16.6 | <0.001 | 0.024 | 81549 | 81649 | 3.1 | 0.379 | 0.036 |
| Model 3C | 87158 | 87342 | 421.2 | <0.001 | 0.040 | 96250 | 96437 | 534.9 | <0.001 | 0.042 | 79241 | 79424 | 2327.9 | <0.001 | 0.136 |
| **Executive Function** | | | | | | | | | | | | | | | |
| Model 1 | 128575 | 128610 |  |  | 0.008 | 75619 | 75652 |  |  | 0.051 | 77447 | 77480 |  |  | 0.034 |
| Model 2A | 128555 | 128599 | 21.8 | <0.001 | 0.008 | 75589 | 75630 | 31.9 | <0.001 | 0.053 | 77437 | 77478 | 11.6 | <0.001 | 0.034 |
| Model 2B | 128526 | 128580 | 30.5 | <0.001 | 0.009 | 75589 | 75639 | 1.4 | 0.240 | 0.054 | 77413 | 77462 | 26.2 | <0.001 | 0.035 |
| Model 3B | 128456 | 128536 | 76.2 | <0.001 | 0.012 | 75403 | 75477 | 192.9 | <0.001 | 0.069 | 77394 | 77469 | 24.3 | <0.001 | 0.036 |
| Model 3A | 128403 | 128509 | 59.3 | <0.001 | 0.012 | 75378 | 75477 | 30.7 | <0.001 | 0.071 | 77395 | 77495 | 5.1 | 0.163 | 0.036 |
| Model 3C | 125005 | 125199 | 3477.7 | <0.001 | 0.126 | 73711 | 73893 | 1717.5 | <0.001 | 0.162 | 75579 | 75761 | 1836.2 | <0.001 | 0.101 |

Model 1: Cognition ~ Age + (1|Participant)

Model 2A: Cognition ~ PP + Age + (1|Participant)

Model 2B: Cognition ~ Spline (PP) + Age + (1|Participant)

Model 3A: Cognition ~ Spline (PP baseline) * Spline (PP change) + Age + (1|Participant)

Model 3B: Cognition ~ Spline (PP baseline) * PP change + Age + (1|Participant)

Model 3C: Cognition ~ Spline (PP baseline) * PP change + Age + Covariates + (1|Participant)

Nonlinear effects of PP were modelled using a restricted cubic spline with three knots.

**Table S9: Model fit and comparison for Restricted Cubic Spline Models with 3–5 Knots**

| **Knot** | **HRS** | | | **ELSA** | | | **CHARLS** | | |
| --- | --- | --- | --- | --- | --- | --- | --- | --- | --- |
|  | **AIC** | **Chisq** | **P** | **AIC** | **Chisq** | **P** | **AIC** | **Chisq** | **P** |
| **Model 2B:** Cognition ~ Spline (PP) + Age + (1\|Participant) | | | | | | | | | |
| **Memory** |  |  |  |  |  |  |  |  |  |
| 3 | 131662 |  |  | 94292 |  |  | 98102 |  |  |
| 4 | 131665 | 0.0 | - | 94294 | 0.0 | - | 98101 | 3.3 | 0.070 |
| 5 | 131666 | 0.8 | 0.357 | 94295 | 0.5 | 0.480 | 98103 | 0.0 | - |
| **Orientation** |  |  |  |  |  |  |  |  |  |
| 3 | 87679 |  |  | 96661 |  |  | 78979 |  |  |
| 4 | 87673 | 7.8 | 0.005 | 96662 | 1.3 | 0.263 | 78981 | 0.0 | - |
| 5 | 87674 | 0.6 | 0.433 | 96660 | 3.2 | 0.072 | 78981 | 2.2 | 0.136 |
| **Executive Function** |  |  |  |  |  |  |  |  |  |
| 3 | 128526 |  |  | 75201 |  |  | 74810 |  |  |
| 4 | 128528 | 0.9 | 0.336 | 75202 | 0.7 | 0.389 | 74810 | 1.5 | 0.214 |
| 5 | 128529 | 0.4 | 0.505 | 75202 | 2.6 | 0.110 | 74812 | 0.0 | - |
| **Model 3B:** Cognition ~ Spline (PP baseline) * PP change + Age + (1\|Participant) | | | | | | | | | |
| **Memory** |  |  |  |  |  |  |  |  |  |
| 3 | 131510 |  |  | 94080 |  |  | 98089 |  |  |
| 4 | 131503 | 10.7 | 0.004 | 94066 | 17.9 | <0.001 | 98093 | 0.7 | 0.718 |
| 5 | 131502 | 5.0 | 0.081 | 94067 | 2.6 | 0.271 | 98096 | 0.8 | 0.684 |
| **Orientation** |  |  |  |  |  |  |  |  |  |
| 3 | 87671 |  |  | 96623 |  |  | 78970 |  |  |
| 4 | 87675 | 0.0 | - | 96624 | 3.4 | 0.181 | 78973 | 0.5 | 0.760 |
| 5 | 87678 | 1.2 | 0.541 | 96618 | 9.5 | 0.008 | 78975 | 2.6 | 0.278 |
| **Executive Function** | | | | | | | | | |
| 3 | 128456 |  |  | 75007 |  |  | 74796 |  |  |
| 4 | 128457 | 3.3 | 0.193 | 74991 | 20.0 | <0.001 | 74799 | 1.7 | 0.428 |
| 5 | 128459 | 2.5 | 0.288 | 74990 | 5.7 | 0.057 | 74801 | 1.3 | 0.510 |

CHARLS=China Health and Retirement Longitudinal Study. HRS=**Health and Retirement Study.** ELSA=**English Longitudinal Study of Ageing. M**odels were adjusted for baseline age, sex, mean arterial pressure, early life factor (education), midlife factors (hearing loss, depression, smoking, drinking, diabetes, hypertension, obesity, and physical inactivity), and late life factors (marital status and visual loss).

**Table S10: Baseline characteristics of participants in different PP groups in HRS**

| **Characteristics** | **Low** | **Normal** | **Borderline** | **High** |
| --- | --- | --- | --- | --- |
| N, % | 1867 (8.2%) | 7394 (32.5%) | 5719 (25.1%) | 7797 (34.2%) |
| Age, mean (SD), year | 55.9 (8.61) | 59.2 (9.25) | 64.1 (9.94) | 69.1 (10.0) |
| Sex, male, % | 290 (15.5%) | 2828 (38.2%) | 2673 (46.7%) | 3688 (47.3%) |
| Ethnicity, white, % | 1278 (68.5%) | 4937 (66.8%) | 4107 (71.8%) | 5663 (72.6%) |
| SBP, mean (SD), mmHg | 107 (13.5) | 119 (12.4) | 129 (14.5) | 144 (21.0) |
| PP, mean (SD), mmHg | 29.8 (6.48) | 40.2 (5.32) | 48.8 (7.55) | 61.7 (14.0) |
| MAP, mean (SD), mmHg | 87.1 (11.7) | 92.2 (11.0) | 96.6 (12.0) | 103 (14.6) |
| Modifiable factors (%) | | | | |
| Education |  |  |  |  |
| None and primary | 61 (3.3%) | 327 (4.4%) | 292 (5.1%) | 519 (6.7%) |
| Secondly | 1794 (96.1%) | 7034 (95.1%) | 5403 (94.5%) | 7256 (93.1%) |
| Marital status, unmarried | 574 (30.7%) | 2296 (31.1%) | 1767 (30.9%) | 2673 (34.3%) |
| Smoking | 1013 (54.3%) | 3940 (53.3%) | 3254 (56.9%) | 4449 (57.1%) |
| Drinking | 1155 (61.9%) | 4602 (62.2%) | 3374 (59.0%) | 4145 (53.2%) |
| Hypertension | 665 (35.6%) | 3230 (43.7%) | 3155 (55.2%) | 5313 (68.1%) |
| Diabetes | 261 (14.0%) | 1205 (16.3%) | 1110 (19.4%) | 2067 (26.5%) |
| Obesity | 965 (51.7%) | 4029 (54.5%) | 3141 (54.9%) | 4234 (54.3%) |
| Hearing loss | 228 (12.2%) | 1135 (15.4%) | 1074 (18.8%) | 1734 (22.2%) |
| Visual loss | 414 (22.2%) | 1684 (22.8%) | 1230 (21.5%) | 1929 (24.7%) |
| Depression | 376 (20.1%) | 1200 (16.2%) | 829 (14.5%) | 1096 (14.1%) |
| Physical inactivity | 91 (4.9%) | 374 (5.1%) | 327 (5.7%) | 551 (7.1%) |
| HRS=**Health and Retirement Study.** SD=standard deviation. SBP=systolic blood pressure. PP=pulse pressure. MAP=mean arterial pressure. | | | | |

**Table S11: Baseline characteristics of participants in different PP groups in ELSA**

| **Characteristics** | **Low** | **Normal** | **Borderline** | **High** |
| --- | --- | --- | --- | --- |
| N, % | 80 (0.6%) | 1898 (14.4%) | 3038 (23.1%) | 8163 (61.9%) |
| Age, mean (SD), year | 56.3 (6.45) | 56.5 (6.65) | 58.9 (8.06) | 65.5 (9.61) |
| Sex, male, % | 9 (11.3%) | 713 (37.6%) | 1500 (49.4%) | 3626 (44.4%) |
| Ethnicity, white, % | 64 (80.0%) | 1586 (83.6%) | 2278 (75.0%) | 5236 (64.1%) |
| SBP, mean (SD), mmHg | 109 (12.4) | 117 (11.2) | 127 (12.5) | 144 (18.8) |
| PP, mean (SD), mmHg | 33.4 (8.61) | 42.6 (4.67) | 51.0 (5.89) | 66.8 (14.1) |
| MAP, mean (SD), mmHg | 87.1 (10.2) | 89.1 (10.4) | 93.2 (11.2) | 99.6 (13.1) |
| Modifiable factors (%) | | | | |
| Education |  |  |  |  |
| None and primary | 22 (27.5%) | 403 (21.2%) | 880 (29.0%) | 3424 (41.9%) |
| Secondly | 57 (71.3%) | 1456 (76.7%) | 2139 (70.4%) | 4726 (57.9%) |
| Marital status, unmarried | 44 (55.0%) | 678 (35.7%) | 986 (32.5%) | 2772 (34.0%) |
| Smoking | 29 (36.3%) | 731 (38.5%) | 1360 (44.8%) | 4236 (51.9%) |
| Drinking | 65 (81.3%) | 1576 (83.0%) | 2627 (86.5%) | 7032 (86.1%) |
| Hypertension | 13 (16.3%) | 316 (16.6%) | 728 (24.0%) | 3378 (41.4%) |
| Diabetes | 3 (3.8%) | 74 (3.9%) | 130 (4.3%) | 674 (8.3%) |
| Obesity | 33 (41.3%) | 501 (26.4%) | 1115 (36.7%) | 3473 (42.5%) |
| Hearing loss | 8 (10.0%) | 253 (13.3%) | 499 (16.4%) | 1676 (20.5%) |
| Visual loss | 13 (16.3%) | 185 (9.7%) | 326 (10.7%) | 1150 (14.1%) |
| Depression | 20 (25.0%) | 288 (15.2%) | 436 (14.4%) | 1237 (15.2%) |
| Physical inactivity | 20 (25.0%) | 216 (11.4%) | 326 (10.7%) | 1319 (16.2%) |
| ELSA=**English Longitudinal Study of Ageing.** SD=deviation. SBP=systolic blood pressure. PP=pulse pressure. MAP=mean arterial pressure. | | | | |

**Table S12: Baseline characteristics of participants in different PP groups in CHARLS**

| **Characteristics** | **Low** | **Normal** | **Borderline** | **High** |
| --- | --- | --- | --- | --- |
| N, % | 275 (1.5%) | 6709 (35.5%) | 5215 (27.6%) | 6723 (35.5%) |
| Age, mean (SD), year | 52.8 (8.29) | 52.6 (7.77) | 56.6 (8.73) | 63.1 (9.65) |
| Sex, male, % | 100 (36.4%) | 3003 (44.8%) | 2708 (51.9%) | 3089 (45.9%) |
| SBP, mean (SD), mmHg | 107 (12.7) | 114 (11.9) | 126 (13.8) | 144 (21.2) |
| PP, mean (SD), mmHg | 32.5 (8.01) | 41.7 (4.76) | 50.6 (6.14) | 65.7 (15.0) |
| MAP, mean (SD), mmHg | 85.0 (11.7) | 86.7 (10.9) | 92.4 (12.2) | 100 (14.5) |
| Modifiable factors (%) | | | | |
| Education |  |  |  |  |
| None and primary | 136 (49.5%) | 2812 (41.9%) | 2802 (53.7%) | 4602 (68.5%) |
| Secondly | 78 (28.4%) | 2053 (30.6%) | 1651 (31.7%) | 1515 (22.5%) |
| Marital status, unmarried | 14 (5.1%) | 398 (5.9%) | 456 (8.7%) | 1107 (16.5%) |
| Smoking | 73 (26.5%) | 2044 (30.5%) | 1905 (36.5%) | 2334 (34.7%) |
| Drinking | 85 (30.9%) | 2428 (36.2%) | 2016 (38.7%) | 2113 (31.4%) |
| Hypertension | 17 (6.2%) | 522 (7.8%) | 863 (16.5%) | 2374 (35.3%) |
| Diabetes | 3 (1.1%) | 199 (3.0%) | 234 (4.5%) | 479 (7.1%) |
| Obesity | 24 (8.7%) | 711 (10.6%) | 688 (13.2%) | 935 (13.9%) |
| Hearing loss | 12 (4.4%) | 370 (5.5%) | 345 (6.6%) | 599 (8.9%) |
| Visual loss | 8 (2.9%) | 270 (4.0%) | 273 (5.2%) | 504 (7.5%) |
| Depression | 110 (40.0%) | 2357 (35.1%) | 1804 (34.6%) | 2495 (37.1%) |
| Physical inactivity | 13 (4.7%) | 251 (3.7%) | 173 (3.3%) | 268 (4.0%) |
| CHARLS=China Health and Retirement Longitudinal Study. SD=standard deviation. SBP=systolic blood pressure. PP=pulse pressure. MAP=mean arterial pressure. | | | | |

**Table S13: Associations of PP thresholds and frequency of PP wave exceedance with cognition**

|  | **HRS** | | **ELSA** | | **CHARLS** | | **Pool** | |
| --- | --- | --- | --- | --- | --- | --- | --- | --- |
|  | **Main Effect** | **Time Interaction Effect** | **Main Effect** | **Time Interaction Effect** | **Main Effect** | **Time Interaction Effect** | **Main Effect** | **Time Interaction Effect** |
| Memory | | | | | | | | |
| PP Group | | | | | | | | |
| Low | -0.077(-0.117, -0.038) ^**^ | 0.002(-0.003, 0.007) | -0.198 (-0.356, -0.039) ^*^ | 0.005 (-0.008, 0.018) | 0.032 (-0.057, 0.120) | -0.003 (-0.020, 0.013) | -0.068 (-0.180, 0.045) | 0.002 (-0.002, 0.007) |
| Normal | Ref | Ref | Ref | Ref | Ref | Ref | Ref | Ref |
| Borderline |  |  |  |  |  |  |  |  |
| 1 wave | -0.042(-0.072, -0.011) ^**^ | -0.014(-0.017, -0.010) ^**^ | 0.004 (-0.044, 0.052) | -0.000 (-0.005, 0.004) | -0.003 (-0.033, 0.028) | -0.010(-0.016, -0.005) ^**^ | -0.016 (-0.045, 0.013) | -0.008 (-0.016, -0.000) ^*^ |
| ≥ 2 waves | -0.025 (-0.068, 0.018) | -0.019(-0.023, -0.014) ^**^ | 0.021 (-0.034, 0.076) | -0.005 (-0.009, -0.001) ^*^ | -0.011 (-0.054, 0.033) | -0.019(-0.026, -0.012) ^**^ | -0.009 (-0.035, 0.018) | -0.014(-0.023, -0.005) ^**^ |
| High |  |  |  |  |  |  |  |  |
| 1 wave | -0.082(-0.114, -0.051) ^**^ | -0.025(-0.029, -0.022) ^**^ | 0.005 (-0.041, 0.050) | -0.009(-0.013, -0.006) ^**^ | -0.069 (-0.103, -0.036 ^**^ | -0.025(-0.030, -0.019) ^**^ | -0.051 (-0.102, -0.000) ^*^ | -0.020(-0.030, -0.009) ^**^ |
| ≥ 2 waves | -0.113(-0.152, -0.074) ^**^ | -0.032(-0.035, -0.028) ^**^ | -0.021 (-0.070, 0.028) | -0.018(-0.022, -0.015) ^**^ | -0.073(-0.114, -0.032)^**^ | -0.045(-0.050, -0.039) ^**^ | -0.071(-0.122, -0.020) ^**^ | -0.031(-0.046, -0.017) ^**^ |
| Orientation | | | | | | | | |
| PP group | | | | | | | | |
| Low | -0.059 (-0.108, -0.009) ^*^ | 0.001 (-0.008, 0.010) | -0.119 (-0.234, -0.005) | 0.004 (-0.011, 0.020) | 0.027 (-0.068, 0.122) | -0.006 (-0.022, 0.011) | -0.048 (-0.116, 0.020) | 0.001 (-0.006, 0.008) |
| Normal | Ref | Ref | Ref | Ref | Ref | Ref | Ref | Ref |
| Borderline |  |  |  |  |  |  |  |  |
| 1 wave | 0.008 (-0.025, 0.041) | -0.011(-0.017, -0.005) ^**^ | 0.008 (-0.028, 0.044) | 0.003 (-0.002, 0.008) | 0.028 (-0.004, 0.061) | -0.001 (-0.006, 0.005) | 0.015 (-0.004, 0.035) | -0.003 (-0.011, 0.005) |
| ≥ 2 waves | 0.014 (-0.027, 0.056) | -0.015(-0.022, -0.008) ^**^ | 0.018 (-0.021, 0.058) | -0.000 (-0.005, 0.004) | -0.004 (-0.051, 0.042) | 0.003 (-0.004, 0.010) | 0.011 (-0.014, 0.035) | -0.004 (-0.014, 0.007) |
| High |  |  |  |  |  |  |  |  |
| 1 wave | -0.005 (-0.038, 0.027) | -0.022(-0.028, -0.017)^**^ | 0.010 (-0.024, 0.044) | -0.002 (-0.007, 0.002) | -0.036 (-0.073, -0.000) ^*^ | -0.005 (-0.010, 0.001) | -0.010 (-0.035, 0.016) | -0.010 (-0.022, 0.002) |
| ≥ 2 waves | -0.016 (-0.053, 0.021) | -0.022(-0.028, -0.016)^**^ | 0.022 (-0.014, 0.058) | -0.009(-0.013, -0.005) ^**^ | -0.045(-0.089, 0.000) ^*^ | -0.011(-0.017, -0.005) ^**^ | -0.011 (-0.048, 0.026) | -0.014(-0.022, -0.006) ^**^ |
| Executive Function | | | | | | | | |
| PP group | | | | | | | | |
| Low | -0.020 (-0.061, 0.021) | 0.002 (-0.002, 0.006) | -0.117 (-0.289, 0.055) | 0.000 (-0.013, 0.014) | 0.009 (-0.081, 0.099) | 0.014 (-0.003, 0.032) | -0.020 (-0.056, 0.017) | 0.003 (-0.001, 0.007) |
| Normal | Ref | Ref | Ref | Ref | Ref | Ref | Ref | Ref |
| Borderline |  |  |  |  |  |  |  |  |
| 1 wave | -0.054(-0.086, -0.022) ^**^ | -0.008(-0.011, -0.005) ^**^ | 0.014 (-0.037, 0.066) | 0.000 (-0.004, 0.005) | 0.022 (-0.008, 0.053) | 0.003 (-0.002, 0.009) | -0.007 (-0.057, 0.043) | -0.002 (-0.009, 0.005) |
| ≥ 2 waves | -0.059(-0.104, -0.014) ^**^ | -0.009(-0.012, -0.005) ^**^ | -0.023 (-0.083, 0.037) | -0.008(-0.013, -0.004) ^**^ | -0.007 (-0.050, 0.036) | 0.001 (-0.006, 0.008) | -0.030 (-0.064, 0.004) | -0.006 (-0.012, -0.001) ^*^ |
| High |  |  |  |  |  |  |  |  |
| 1 wave | -0.083(-0.117, -0.050) ^**^ | -0.016(-0.019, -0.013) ^**^ | -0.020 (-0.068, 0.029) | -0.011(-0.015, -0.007) ^**^ | 0.005 (-0.029, 0.040) | 0.001 (-0.005, 0.007) | -0.033 (-0.087, 0.021) | -0.009 (-0.019, 0.001) |
| ≥ 2 waves | -0.114(-0.155, -0.073) ^**^ | -0.018(-0.021, -0.015) ^**^ | -0.024 (-0.077, 0.029) | -0.017(-0.021, -0.014) ^**^ | -0.037 (-0.079, 0.005) | 0.003 (-0.003, 0.009) | -0.060 (-0.116, -0.004) ^*^ | -0.011 (-0.025, 0.002) |
| * p < 0.05. ** p < 0.05/3 (Bonferroni correction). CHARLS=China Health and Retirement Longitudinal Study. HRS=**Health and Retirement Study.** ELSA=**English Longitudinal Study of Ageing.** PP=pulse pressure. Models were adjusted for baseline age, sex, mean arterial pressure, early life factor (education), midlife factors (hearing loss, depression, smoking, drinking, diabetes, hypertension, obesity, and physical inactivity), and late life factors (marital status and visual loss). | | | | | | | | |

**Table S14: Associations of PP thresholds with cognition considering antihypertensive medication use**

|  | **HRS** | | **ELSA** | | **CHARLS** | | **Pool** | |
| --- | --- | --- | --- | --- | --- | --- | --- | --- |
|  | **Main Effect** | **Time Interaction Effect** | **Main Effect** | **Time Interaction Effect** | **Main Effect** | **Time Interaction Effect** | **Main Effect** | **Time Interaction Effect** |
| Memory | | | | | | | | |
| PP Group | | | | | | | | |
| Low | -0.080(-0.120, -0.041) ^**^ | 0.002 (-0.003, 0.008) | -0.199(-0.357, -0.040)^**^ | 0.004 (-0.009, 0.017) | 0.032 (-0.057, 0.120) | -0.003 (-0.020, 0.013) | -0.070 (-0.183, 0.044) | 0.002 (-0.002, 0.007) |
| Normal | Ref | Ref | Ref | Ref | Ref | Ref | Ref | Ref |
| Borderline | -0.039(-0.067, -0.011) ^**^ | -0.015(-0.019, -0.012) ^**^ | 0.004 (-0.041, 0.048) | -0.003 (-0.006, 0.001) | -0.006 (-0.034, 0.022) | -0.013(-0.018, -0.008) ^**^ | -0.017 (-0.042, 0.009) | -0.010(-0.018, -0.003) ^**^ |
| High | -0.095(-0.125, -0.065)^**^ | -0.028(-0.031, -0.025) ^**^ | -0.015 (-0.060, 0.029) | -0.015(-0.019, -0.012) ^**^ | -0.073(-0.105, -0.041) ^**^ | -0.034(-0.038, -0.029) ^**^ | -0.064 (-0.108, -0.019) ^*^ | -0.026(-0.036, -0.015) ^**^ |
| Orientation | | | | | | | | |
| PP group | | | | | | | | |
| Low | -0.059 (-0.108, -0.009)^*^ | 0.002 (-0.007, 0.011) | -0.124 (-0.238, -0.010) ^*^ | 0.004 (-0.011, 0.019) | 0.028 (-0.067, 0.123) | -0.006 (-0.023, 0.011) | -0.049 (-0.120, 0.023) | 0.001 (-0.006, 0.008) |
| Normal | Ref | Ref | Ref | Ref | Ref | Ref | Ref | Ref |
| Borderline | 0.008 (-0.022, 0.038) | -0.013(-0.018, -0.007) ^**^ | 0.005 (-0.028, 0.038) | 0.001 (-0.004, 0.005) | 0.020 (-0.010, 0.050) | 0.001 (-0.004, 0.006) | 0.011 (-0.006, 0.029) | -0.004 (-0.012, 0.005) |
| High | -0.011 (-0.041, 0.020) | -0.022(-0.027, -0.017) ^**^ | 0.004 (-0.029, 0.037) | -0.006(-0.010, -0.002) ^**^ | -0.039 (-0.073, -0.004) ^*^ | -0.007(-0.012, -0.003) ^**^ | -0.015 (-0.038, 0.009) | -0.012(-0.021, -0.002) ^**^ |
| Executive Function | | | | | | | | |
| PP group | | | | | | | | |
| Low | -0.020 (-0.061, 0.021) | 0.002 (-0.002, 0.006) | -0.113 (-0.284, 0.059) | 0.000 (-0.013, 0.014) | 0.010 (-0.080, 0.100) | 0.014 (-0.004, 0.032) | -0.019 (-0.056, 0.017) | 0.003 (-0.001, 0.006) |
| Normal | Ref | Ref | Ref | Ref | Ref | Ref | Ref | Ref |
| Borderline | -0.055(-0.084, -0.026) ^**^ | -0.009(-0.011, -0.006) ^**^ | -0.002 (-0.050, 0.046) | -0.004 (-0.008, 0.000) | 0.017 (-0.011, 0.045) | 0.002 (-0.003, 0.008) | -0.014 (-0.059, 0.031) | -0.004 (-0.010, 0.003) |
| High | -0.094(-0.125, -0.063) ^**^ | -0.017(-0.020, -0.015) ^**^ | -0.020 (-0.068, 0.028) | -0.015(-0.018, -0.011) ^**^ | -0.006 (-0.039, 0.026) | 0.002 (-0.003, 0.007) | -0.041 (-0.096, 0.014) | -0.010 (-0.022, 0.001) |
| * p < 0.05. ** p < 0.05/3 (Bonferroni correction). CHARLS=China Health and Retirement Longitudinal Study. HRS=**Health and Retirement Study.**ELSA=**English Longitudinal Study of Ageing.** PP=pulse pressure. Models were adjusted for baseline age, sex, mean arterial pressure, early life factor (education), midlife factors (hearing loss, depression, smoking, drinking, diabetes, hypertension, obesity, and physical inactivity), late life factors (marital status and visual loss), and antihypertensive medication. | | | | | | | | |

**Table S15: Associations of PP thresholds with cognition among participants with PP follow-up assessments**

|  | **HRS** | | **ELSA** | | **CHARLS** | | **Pool** | |
| --- | --- | --- | --- | --- | --- | --- | --- | --- |
|  | **Main Effect** | **Time Interaction Effect** | **Main Effect** | **Time Interaction Effect** | **Main Effect** | **Time Interaction Effect** | **Main Effect** | **Time Interaction Effect** |
| Memory | | | | | | | | |
| PP Group | | | | | | | | |
| Low | -0.063(-0.109, -0.017) ^**^ | 0.002 (-0.004, 0.007) | -0.220 (-0.401, -0.039) ^*^ | 0.004 (-0.010, 0.017) | 0.025 (-0.078, 0.127) | -0.001 (-0.019, 0.017) | -0.066 (-0.176, 0.044) | 0.002 (-0.003, 0.006) |
| Normal | Ref | Ref | Ref | Ref | Ref | Ref | Ref | Ref |
| Borderline | -0.063 (-0.109, -0.017) | -0.015(-0.019, -0.012) ^**^ | 0.024 (-0.030, 0.077) | -0.002 (-0.006, 0.002) | 0.015 (-0.017, 0.048) | -0.010(-0.016, -0.004) ^**^ | -0.000 (-0.035, 0.035) | -0.009 (-0.017, -0.001) ^*^ |
| High | -0.084(-0.117, -0.050)^**^ | -0.029(-0.032, -0.026) ^**^ | 0.011 (-0.041, 0.064) | -0.015(-0.018, -0.011) ^**^ | -0.056(-0.091, -0.020) ^**^ | -0.029(-0.034, -0.024) ^**^ | -0.046 (-0.098, 0.007) | -0.024(-0.033, -0.015) ^**^ |
| Orientation | | | | | | | | |
| PP group | | | | | | | | |
| Low | -0.061 (-0.116, -0.006) ^*^ | 0.002 (-0.008, 0.012) | -0.091 (-0.217, 0.036) | 0.002 (-0.015, 0.018) | 0.044 (-0.070, 0.158) | -0.006 (-0.025, 0.013) | -0.042 (-0.106, 0.021) | 0.000 (-0.007, 0.008) |
| Normal | Ref | Ref | Ref | Ref | Ref | Ref | Ref | Ref |
| Borderline | 0.002 (-0.032, 0.035) | -0.011(-0.017, -0.005) ^**^ | 0.023 (-0.016, 0.061) | 0.000 (-0.005, 0.005) | 0.012 (-0.024, 0.049) | 0.001 (-0.005, 0.007) | 0.011 (-0.009, 0.032) | -0.003 (-0.011, 0.005) |
| High | -0.013 (-0.047, 0.020) | -0.021(-0.026, -0.015) ^**^ | 0.034 (-0.003, 0.071) | -0.008(-0.012, -0.003) ^**^ | -0.046 (-0.086, -0.006)^*^ | -0.008(-0.014, -0.003) ^**^ | -0.008 (-0.053, 0.037) | -0.012(-0.020, -0.004) ^**^ |
| Executive Function | | | | | | | | |
| PP group | | | | | | | | |
| Low | -0.035 (-0.084, 0.014) | 0.002 (-0.003, 0.006) | -0.058 (-0.254, 0.138) | -0.000 (-0.014, 0.014) | -0.013 (-0.123, 0.097) | 0.008 (-0.012, 0.028) | -0.034 (-0.077, 0.010) | 0.002 (-0.002, 0.006) |
| Normal | Ref | Ref | Ref | Ref | Ref | Ref | Ref | Ref |
| Borderline | -0.077(-0.111, -0.042) ^**^ | -0.009(-0.012, -0.006) ^**^ | 0.004 (-0.054, 0.062) | -0.005 (-0.010, -0.001) ^*^ | 0.011 (-0.023, 0.046) | 0.002 (-0.003, 0.008) | -0.023 (-0.080, 0.035) | -0.004 (-0.011, 0.002) |
| High | -0.117(-0.153, -0.081) ^**^ | -0.018(-0.020, -0.015) ^**^ | -0.022 (-0.079, 0.034) | -0.016(-0.020, -0.012) ^**^ | -0.016 (-0.054, 0.022) | 0.002 (-0.004, 0.008) | -0.054 (-0.119, 0.011) | -0.011 (-0.023, 0.001) |
| * p < 0.05. ** p < 0.05/3 (Bonferroni correction). CHARLS=China Health and Retirement Longitudinal Study. HRS=**Health and Retirement Study.** ELSA=**English Longitudinal Study of Ageing.** PP=pulse pressure. Models were adjusted for baseline age, sex, mean arterial pressure, early life factor (education), midlife factors (hearing loss, depression, smoking, drinking, diabetes, hypertension, obesity, and physical inactivity), and late life factors (marital status and visual loss). | | | | | | | | |

**Table S16: Associations of PP thresholds with cognition among participants based on pattern‑mixture models**

| **PP Group** | **HRS** | | **ELSA** | | **CHARLS** | | **Pool** | |
| --- | --- | --- | --- | --- | --- | --- | --- | --- |
|  | **Main Effect** | **Time Interaction Effect** | **Main Effect** | **Time Interaction Effect** | **Main Effect** | **Time Interaction Effect** | **Main Effect** | **Time Interaction Effect** |
| **Memory** | | | | | | | | |
| ≤ 2 Visits | | | | | | | | |
| Low | -0.061 (-0.116, -0.006) ^*^ | 0.011 (0.002, 0.021) ^*^ | -0.184 (-0.438, 0.069) | 0.012 (-0.016, 0.040) | 0.073 (-0.044, 0.191) | 0.022 (-0.002, 0.046) | -0.032 (-0.149, 0.084) | 0.013 (0.004, 0.021) ^**^ |
| Normal | Ref | Ref | Ref | Ref | Ref | Ref | Ref | Ref |
| Borderline | -0.035 (-0.074, 0.004) | -0.018 (-0.024, -0.011) ^**^ | 0.020 (-0.039, 0.079) | 0.004 (-0.003, 0.010) | -0.011 (-0.047, 0.025) | -0.015 (-0.022, -0.009) ^**^ | -0.015 (-0.040, 0.010) | -0.010 (-0.023, 0.004) |
| High | -0.111 (-0.153, -0.068) ^**^ | -0.034 (-0.040, -0.028) ^**^ | -0.024 (-0.085, 0.036) | -0.014 (-0.020, -0.008) ^**^ | -0.091 (-0.134, -0.049) ^**^ | -0.038 (-0.045, -0.031) ^**^ | -0.080(-0.127, -0.033) ^**^ | -0.029 (-0.043, -0.014) ^**^ |
| ≥ 3 Visits | | | | | | | | |
| Low | -0.042 (-0.103, 0.019) | -0.002 (-0.008, 0.004) | -0.216 (-0.432, 0.000) ^*^ | 0.003 (-0.012, 0.018) | 0.006 (-0.129, 0.142) | -0.020 (-0.043, 0.003) | -0.045 (-0.099, 0.009) | -0.002 (-0.008, 0.003) |
| Normal | Ref | Ref | Ref | Ref | Ref | Ref | Ref | Ref |
| Borderline | -0.011 (-0.054, 0.033) | -0.015(-0.020, -0.011)^**^ | -0.008 (-0.085, 0.068) | -0.003 (-0.009, 0.002) | 0.003 (-0.042, 0.048) | -0.006 (-0.013, 0.002) | -0.005 (-0.034, 0.024) | -0.008 (-0.016, -0.001) ^*^ |
| High | -0.052(-0.097, -0.008) ^*^ | -0.028(-0.032, -0.024) ^**^ | -0.002 (-0.076, 0.072) | -0.015 (-0.019, -0.010) ^**^ | -0.053 (-0.101, -0.005) ^*^ | -0.024 (-0.031, -0.017) ^**^ | -0.044(-0.074, -0.015) ^**^ | -0.022 (-0.030, -0.014) ^**^ |
| Overall effect | | | | | | | | |
| Low | -0.052 (-0.093, -0.011)^**^ | 0.004 (-0.009, 0.017) | -0.203(-0.367, -0.038)^**^ | 0.005 (-0.008, 0.018) | 0.045 (-0.044, 0.133) | 0.001 (-0.040, 0.042) | -0.054 (-0.175, 0.066) | 0.004 (-0.005, 0.013) |
| Normal | Ref | Ref | Ref | Ref | Ref | Ref | Ref | Ref |
| Borderline | -0.024 (-0.053, 0.005) | -0.016 (-0.019, -0.012) ^**^ | 0.009 (-0.037, 0.056) | -0.000 (-0.007, 0.007) | -0.005 (-0.034, 0.023) | -0.011 (-0.020, -0.001) ^*^ | -0.011 (-0.029, 0.008) | -0.009 (-0.019, 0.000) |
| High | -0.082 (-0.139, -0.025) | -0.030 (-0.036, -0.024) ^**^ | -0.016 (-0.062, 0.031) | -0.014 (-0.018, -0.011) ^**^ | -0.074 (-0.111, -0.037) ^**^ | -0.031 (-0.045, -0.017) ^**^ | -0.057 (-0.097, -0.016) ^**^ | -0.024 (-0.036, -0.013) ^**^ |
| **Orientation** |  |  |  |  |  |  |  |  |
| ≤ 2 Visits |  |  |  |  |  |  |  |  |
| Low | -0.091 (-0.178, -0.003) ^*^ | -0.009 (-0.027, 0.010) | -0.036 (-0.250, 0.178) | 0.031 (-0.002, 0.065) | 0.056 (-0.065, 0.178) | -0.003 (-0.027, 0.021) | -0.028 (-0.132, 0.075) | 0.003 (-0.018, 0.024) |
| Normal | Ref | Ref | Ref | Ref | Ref | Ref | Ref | Ref |
| Borderline | 0.055 (0.006, 0.104) ^*^ | -0.017 (-0.027, -0.006) ^**^ | -0.007 (-0.057, 0.043) | 0.001 (-0.007, 0.008) | 0.034 (-0.003, 0.072) | -0.006 (-0.012, 0.001) | 0.029 (-0.004, 0.061) | -0.007 (-0.016, 0.003) |
| High | -0.016 (-0.066, 0.033) | -0.042 (-0.052, -0.032) ^**^ | -0.019 (-0.069, 0.032) | -0.009 (-0.016, -0.002)^**^ | -0.039 (-0.083, 0.006) | -0.013 (-0.019, -0.006) ^**^ | -0.026 (-0.054, 0.002) | -0.021 (-0.041, -0.001) ^*^ |
| ≥ 3 Visits |  |  |  |  |  |  |  |  |
| Low | -0.046 (-0.111, 0.020) | 0.003 (-0.008, 0.014) | -0.086 (-0.230, 0.058) | -0.004 (-0.022, 0.014) | -0.015 (-0.168, 0.139) | -0.006 (-0.030, 0.018) | -0.048 (-0.103, 0.008) | 0.000 (-0.009, 0.009) |
| Normal | Ref | Ref | Ref | Ref | Ref | Ref | Ref | Ref |
| Borderline | -0.019 (-0.061, 0.023) | -0.009 (-0.016, -0.002) ^**^ | 0.054 (0.003, 0.105) ^*^ | 0.000 (-0.006, 0.006) | -0.012 (-0.063, 0.040) | 0.007 (-0.000, 0.015) | 0.007 (-0.039, 0.052) | -0.001 (-0.010, 0.009) |
| High | -0.014 (-0.055, 0.028) | -0.018 (-0.024, -0.012) ^**^ | 0.065 (0.016, 0.114) ^**^ | -0.007 (-0.013, -0.002) ^**^ | -0.054 (-0.109, 0.000) | -0.003 (-0.010, 0.004) | -0.001 (-0.068, 0.067) | -0.010 (-0.018, -0.001) ^*^ |
| Overall effect |  |  |  |  |  |  |  |  |
| Low | -0.062 (-0.114, -0.009) ^*^ | -0.000 (-0.011, 0.010) | -0.070 (-0.190, 0.049) | 0.011 (-0.023, 0.045) | 0.029 (-0.067, 0.124) | -0.004 (-0.021, 0.013) | -0.039 (-0.097, 0.018) | -0.001 (-0.009, 0.008) |
| Normal | Ref | Ref | Ref | Ref | Ref | Ref | Ref | Ref |
| Borderline | 0.017 (-0.056, 0.090) | -0.012 (-0.019, -0.005) ^**^ | 0.023 (-0.037, 0.084) | 0.000 (-0.004, 0.005) | 0.015 (-0.030, 0.060) | 0.001 (-0.012, 0.013) | 0.018 (-0.015, 0.050) | -0.004 (-0.013, 0.005) |
| High | -0.015 (-0.047, 0.017) | -0.030 (-0.053, -0.006) ^**^ | 0.023 (-0.059, 0.105) | -0.008 (-0.012, -0.004) ^**^ | -0.045 (-0.079, -0.010) ^**^ | -0.008 (-0.018, 0.002) | -0.023 (-0.052, 0.005) | -0.009 (-0.013, -0.005) ^**^ |
| **Executive Function** | | | | | | | | |
| ≤ 2 Visits |  |  |  |  |  |  |  |  |
| Low | -0.004 (-0.060, 0.053) | 0.007 (-0.001, 0.015) | -0.024 (-0.292, 0.244) | 0.025 (-0.006, 0.056) | 0.089 (-0.021, 0.199) | 0.017 (-0.007, 0.040) | 0.024 (-0.072, 0.120) | 0.009 (0.001, 0.017) ^*^ |
| Normal | Ref | Ref | Ref | Ref | Ref | Ref | Ref | Ref |
| Borderline | -0.039 (-0.079, 0.000) | -0.011 (-0.017, -0.006) ^**^ | 0.014 (-0.048, 0.077) | 0.002 (-0.005, 0.008) | 0.021 (-0.013, 0.055) | 0.002 (-0.005, 0.009) | 0.002 (-0.028, 0.033) | -0.003 (-0.012, 0.006) |
| High | -0.087 (-0.130, -0.044) ^**^ | -0.024 (-0.029, -0.019) ^**^ | -0.045 (-0.109, 0.018) | -0.015 (-0.021, -0.009) ^**^ | 0.002 (-0.039, 0.042) | 0.005 (-0.002, 0.012) | -0.032 (-0.071, 0.008) | -0.011 (-0.028, 0.006) |
| ≥ 3 Visits |  |  |  |  |  |  |  |  |
| Low | -0.020 (-0.084, 0.044) | 0.000 (-0.005, 0.005) | -0.167 (-0.403, 0.070) | -0.007 (-0.022, 0.009) | -0.115 (-0.268, 0.039) | 0.013 (-0.013, 0.040) | -0.060 (-0.131, 0.012) | -0.000 (-0.005, 0.005) |
| Normal | Ref | Ref | Ref | Ref | Ref | Ref | Ref | Ref |
| Borderline | -0.064 (-0.110, -0.018) ^**^ | -0.008 (-0.012, -0.005) ^**^ | -0.027 (-0.111, 0.056) | -0.007 (-0.013, -0.002) ^**^ | 0.002 (-0.048, 0.051) | 0.002 (-0.005, 0.010) | -0.025 (-0.061, 0.011) | -0.005 (-0.011, 0.001) |
| High | -0.097 (-0.144, -0.050) ^**^ | -0.017 (-0.020, -0.013) ^**^ | -0.011 (-0.092, 0.070) | -0.017 (-0.022, -0.013) ^**^ | -0.023 (-0.077, 0.030) | -0.001 (-0.008, 0.007) | -0.040 (-0.075, -0.005) ^*^ | -0.012 (-0.022, -0.002) ^*^ |
| Overall effect | | | | | | | | |
| Low | -0.027 (-0.068, 0.014) | 0.003 (-0.004, 0.009) | -0.104 (-0.281, 0.073) | 0.006 (-0.024, 0.036) | -0.005 (-0.204, 0.193) | 0.015 (-0.003, 0.033) | -0.029 (-0.068, 0.010) | 0.005 (-0.003, 0.012) |
| Normal | Ref | Ref | Ref | Ref | Ref | Ref | Ref | Ref |
| Borderline | -0.033 (-0.062, -0.004) ^*^ | -0.009 (-0.012, -0.006) ^**^ | -0.001 (-0.051, 0.049) | -0.003 (-0.012, 0.006) | 0.015 (-0.013, 0.043) | 0.002 (-0.004, 0.008) | -0.008 (-0.039, 0.024) | -0.004 (-0.011, 0.004) |
| High | -0.060 (-0.091, -0.030) ^**^ | -0.020 (-0.027, -0.013) ^**^ | -0.032 (-0.082, 0.018) | -0.016 (-0.020, -0.013) ^**^ | -0.008 (-0.040, 0.025) | 0.002 (-0.004, 0.008) | -0.034 (-0.068, 0.000) | -0.011 (-0.025, 0.002) |
| * p < 0.05. ** p < 0.05/3 (Bonferroni correction). CHARLS=China Health and Retirement Longitudinal Study. HRS=**Health and Retirement Study.** ELSA=**English Longitudinal Study of Ageing.** PP=pulse pressure. Models were adjusted for baseline age, sex, mean arterial pressure, early life factor (education), midlife factors (hearing loss, depression, smoking, drinking, diabetes, hypertension, obesity, and physical inactivity), and late life factors (marital status and visual loss). | | | | | | | | |

**Table S17: Associations of PP thresholds after excluding participants with baseline stroke and heart disease**

|  | **HRS** | | **ELSA** | | **CHARLS** | | **Pool** | |
| --- | --- | --- | --- | --- | --- | --- | --- | --- |
|  | **Main Effect** | **Time Interaction Effect** | **Main Effect** | **Time Interaction Effect** | **Main Effect** | **Time Interaction Effect** | **Main Effect** | **Time Interaction Effect** |
| Memory | | | | | | | | |
| PP Group | | | | | | | | |
| Low | -0.065(-0.108, -0.021) ^**^ | 0.002 (-0.004, 0.007) | -0.141 (-0.256, -0.025) ^*^ | 0.008 (-0.005, 0.022) | 0.041 (-0.052, 0.135) | -0.004 (-0.022, 0.013) | -0.065 (-0.193, 0.062) | 0.002 (-0.003, 0.007) |
| Normal | Ref | Ref | Ref | Ref | Ref | Ref | Ref | Ref |
| Borderline | -0.024 (-0.056, 0.007) | -0.016(-0.020, -0.012) ^**^ | 0.010 (-0.023, 0.044) | -0.003 (-0.007, 0.001) | -0.002 (-0.032, 0.027) | -0.014(-0.020, -0.009) ^**^ | -0.010 (-0.029, 0.010) | -0.011(-0.019, -0.003) ^**^ |
| High | -0.098(-0.132, -0.063) ^**^ | -0.028(-0.032, -0.024) ^**^ | 0.018 (-0.015, 0.052) | -0.014(-0.018, -0.011) ^**^ | -0.069(-0.102, -0.035) ^**^ | -0.035(-0.040, -0.030) ^**^ | -0.062(-0.109, -0.014)^**^ | -0.025(-0.037, -0.014) ^**^ |
| Orientation | | | | | | | | |
| PP group | | | | | | | | |
| Low | -0.036 (-0.090, 0.018) | 0.001 (-0.008, 0.011) | -0.141(-0.256, -0.025) ^*^ | 0.004 (-0.012, 0.019) | 0.055 (-0.045, 0.155) | -0.002 (-0.020, 0.015) | -0.037 (-0.136, 0.063) | 0.001 (-0.006, 0.009) |
| Normal | Ref | Ref | Ref | Ref | Ref | Ref | Ref | Ref |
| Borderline | 0.018 (-0.016, 0.051) | -0.011(-0.017, -0.005) ^**^ | 0.010 (-0.023, 0.044) | 0.002 (-0.003, 0.006) | 0.016 (-0.016, 0.048) | 0.001 (-0.005, 0.006) | 0.015 (-0.004, 0.034) | -0.003 (-0.011, 0.005) |
| High | -0.008 (-0.042, 0.027) | -0.020(-0.026, -0.015) ^**^ | 0.018 (-0.015, 0.052) | -0.005(-0.009, -0.001) ^**^ | -0.040 (-0.076, -0.003) ^*^ | -0.007(-0.012, -0.002) ^**^ | -0.009 (-0.042, 0.024) | -0.011(-0.020, -0.002) ^*^ |
| Executive Function | | | | | | | | |
| PP group | | | | | | | | |
| Low | -0.016 (-0.061, 0.029) | 0.002 (-0.003, 0.006) | -0.116 (-0.300, 0.067) | 0.000 (-0.014, 0.014) | 0.023 (-0.072, 0.117) | 0.021 (0.002, 0.039) ^*^ | -0.014 (-0.053, 0.026) | 0.005 (-0.005, 0.015) |
| Normal | Ref | Ref | Ref | Ref | Ref | Ref | Ref | Ref |
| Borderline | -0.038(-0.070, -0.006) ^*^ | -0.008(-0.011, -0.005) ^**^ | -0.116 (-0.300, 0.067) | -0.004 (-0.008, 0.000) | 0.019 (-0.011, 0.049) | 0.003 (-0.002, 0.008) | -0.009 (-0.046, 0.027) | -0.003 (-0.010, 0.003) |
| High | -0.098(-0.134, -0.063) ^**^ | -0.015(-0.017, -0.012) ^**^ | -0.116 (-0.300, 0.067) | -0.014(-0.018, -0.010) ^**^ | -0.008 (-0.042, 0.027) | 0.000 (-0.005, 0.005) | -0.042 (-0.100, 0.016) | -0.010(-0.019, -0.001) ^*^ |
| * p < 0.05. ** p < 0.05/3 (Bonferroni correction). CHARLS=China Health and Retirement Longitudinal Study. HRS=**Health and Retirement Study.** ELSA=**English Longitudinal Study of Ageing.** PP=pulse pressure. Models were adjusted for baseline age, sex, mean arterial pressure, early life factor (education), midlife factors (hearing loss, depression, smoking, drinking, diabetes, hypertension, obesity, and physical inactivity), and late life factors (marital status and visual loss). | | | | | | | | |

**Table S18: Associations of PP thresholds with cognition in complete case analysis**

|  | **HRS** | | **ELSA** | | **CHARLS** | | **Pool** | |
| --- | --- | --- | --- | --- | --- | --- | --- | --- |
|  | **Main Effect** | **Time Interaction Effect** | **Main Effect** | **Time Interaction Effect** | **Main Effect** | **Time Interaction Effect** | **Main Effect** | **Time Interaction Effect** |
| Memory | | | | | | | | |
| PP Group | | | | | | | | |
| Low | -0.085(-0.126, -0.043)^**^ | 0.004 (-0.002, 0.009) | -0.227 (-0.438, -0.017) ^*^ | 0.008 (-0.008, 0.023) | 0.035 (-0.076, 0.145) | 0.002 (-0.017, 0.021) | -0.071 (-0.188, 0.046) | 0.004 (-0.001, 0.008) |
| Normal | Ref | Ref | Ref | Ref | Ref | Ref | Ref | Ref |
| Borderline | -0.036 (-0.065, -0.006) ^*^ | -0.015(-0.019, -0.012) ^**^ | 0.013 (-0.043, 0.069) | -0.004 (-0.009, 0.000) | 0.000 (-0.034, 0.034) | -0.009(-0.015, -0.003) ^**^ | -0.013 (-0.042, 0.017) | -0.010(-0.017, -0.004) ^**^ |
| High | -0.089(-0.120, -0.057) ^**^ | -0.029(-0.032, -0.025) ^**^ | 0.006 (-0.049, 0.061) | -0.018(-0.022, -0.013) ^**^ | -0.063(-0.100, -0.025)^**^ | -0.028(-0.033, -0.022) ^**^ | -0.053 (-0.105, -0.001)^*^ | -0.025(-0.032, -0.018) ^**^ |
| Orientation | | | | | | | | |
| PP group | | | | | | | | |
| Low | -0.058 (-0.109, -0.007) ^*^ | 0.002 (-0.007, 0.012) | -0.040 (-0.191, 0.111) | 0.006 (-0.013, 0.024) | 0.030 (-0.093, 0.153) | -0.011 (-0.031, 0.009) | -0.043 (-0.091, 0.005) | 0.001 (-0.007, 0.008) |
| Normal | Ref | Ref | Ref | Ref | Ref | Ref | Ref | Ref |
| Borderline | 0.012 (-0.019, 0.043) | -0.012(-0.018, -0.006) ^**^ | 0.019 (-0.023, 0.061) | 0.001 (-0.005, 0.006) | 0.001 (-0.037, 0.039) | 0.001 (-0.005, 0.007) | 0.010 (-0.011, 0.031) | -0.003 (-0.012, 0.005) |
| High | -0.005 (-0.037, 0.026) | -0.021(-0.027, -0.017) ^**^ | 0.020 (-0.021, 0.061) | -0.007(-0.011, -0.002) ^**^ | -0.050 (-0.091, -0.008) ^*^ | -0.010(-0.015, -0.004) ^**^ | -0.011 (-0.049, 0.026) | -0.012(-0.021, -0.003) ^**^ |
| Executive Function | | | | | | | | |
| PP group | | | | | | | | |
| Low | -0.022 (-0.065, 0.021) | 0.003 (-0.001, 0.007) | -0.079 (-0.306, 0.148) | 0.015 (-0.001, 0.031) | -0.020 (-0.142, 0.102) | 0.008 (-0.013, 0.029) | -0.023 (-0.063, 0.017) | 0.006 (-0.002, 0.014) |
| Normal | Ref | Ref | Ref | Ref | Ref | Ref | Ref | Ref |
| Borderline | -0.055(-0.085, -0.024) ^**^ | -0.009(-0.012, -0.006) ^**^ | -0.001 (-0.061, 0.059) | -0.006(-0.011, -0.001) ^**^ | 0.020 (-0.017, 0.057) | 0.001 (-0.005, 0.007) | -0.014 (-0.062, 0.034) | -0.005 (-0.011, 0.000) |
| High | -0.095(-0.127, -0.062) ^**^ | -0.017(-0.020, -0.015) ^**^ | -0.014 (-0.073, 0.045) | -0.015(-0.020, -0.011) ^**^ | -0.002 (-0.043, 0.038) | 0.001 (-0.005, 0.006) | -0.039 (-0.099, 0.021) | -0.011 (-0.022, 0.000) |
| * p < 0.05. ** p < 0.05/3 (Bonferroni correction). CHARLS=China Health and Retirement Longitudinal Study. HRS=**Health and Retirement Study.** ELSA=**English Longitudinal Study of Ageing.** PP=pulse pressure. Models were adjusted for baseline age, sex, mean arterial pressure, early life factor (education), midlife factors (hearing loss, depression, smoking, drinking, diabetes, hypertension, obesity, and physical inactivity), and late life factors (marital status and visual loss). | | | | | | | | |

**Table S19: Associations of PP thresholds with cognition in one-step IPD analysis**

|  | **Main Effect** | | **Time Interaction Effect** | |
| --- | --- | --- | --- | --- |
|  | **β (95%CI)** | **p** | **β (95%CI)** | **p** |
| Memory | | | | |
| PP group |  |  |  |  |
| Low | -0.053 (-0.087, -0.019) | 0.002^**^ | -0.007 (-0.011, -0.002) | 0.002^**^ |
| Normal | Ref |  | Ref |  |
| Borderline | -0.025 (-0.043, -0.007) | 0.007^**^ | -0.015 (-0.017, -0.013) | <0.001^**^ |
| High | -0.082 (-0.101, -0.062) | <0.001^**^ | -0.030 (-0.032, -0.028) | <0.001^**^ |
| Orientation | | | | |
| PP group |  |  |  |  |
| Low | -0.014 (-0.055, 0.026) | 0.484 | 0.007 (0.000, 0.014) | 0.042 |
| Normal | Ref |  | Ref |  |
| Borderline | 0.006 (-0.011, 0.024) | 0.490 | 0.005 (0.002, 0.008) | <0.001^**^ |
| High | -0.023 (-0.042, -0.005) | 0.015 | 0.001 (-0.001, 0.004) | 0.242 |
| Executive Function | | | | |
| PP group |  |  |  |  |
| Low | -0.044 (-0.080, -0.009) | 0.015^**^ | 0.004 (0.001, 0.008) | 0.024 |
| Normal | Ref |  | Ref |  |
| Borderline | 0.001 (-0.018, 0.020) | 0.938 | -0.005 (-0.007, -0.003) | <0.001^**^ |
| High | -0.040 (-0.060, -0.019) | <0.001^**^ | -0.013 (-0.015, -0.012) | <0.001^**^ |
| * p < 0.05. ** p < 0.05/3 (Bonferroni correction). PP=pulse pressure. IPD analysis=Individual Patient Data analysis. Models were adjusted for baseline age, sex, mean arterial pressure, early life factor (education), midlife factors (hearing loss, depression, smoking, drinking, diabetes, hypertension, obesity, and physical inactivity), and late life factors (marital status and visual loss). Random intercepts for study and participant were considered. | | | | |

**Table S20: Associations of baseline hypertension, diabetes, and smoking with memory**

|  | **HRS** | | **ELSA** | | **CHARLS** | | **Pool** | |
| --- | --- | --- | --- | --- | --- | --- | --- | --- |
|  | **Main Effect** | **Time Interaction Effect** | **Main Effect** | **Time Interaction Effect** | **Main Effect** | **Time Interaction Effect** | **Main Effect** | **Time Interaction Effect** |
| Hypertension | -0.091(-0.112, -0.069) ^**^ | -0.010 (-0.012, -0.007) ^**^ | -0.018(-0.046, 0.011) ^*^ | -0.007(-0.009, -0.005) ^**^ | 0.039 (0.010, 0.068) ^**^ | -0.011(-0.015, -0.006) ^**^ | -0.072 (-0.087, -0.057) ^**^ | -0.012 (-0.014, -0.011) ^**^ |
| Diabetes | -0.111 (-0.137, -0.085) ^**^ | -0.010 (-0.013, -0.007) ^**^ | -0.143 (-0.194, -0.091) ^**^ | -0.010 (-0.015, -0.006) ^**^ | 0.019 (-0.030, 0.068) | -0.008 (-0.017, 0.001) | -0.133 (-0.154, -0.112) | -0.012 (-0.015, -0.010) ^**^ |
| Smoking | -0.108 (-0.129, -0.088) ^**^ | -0.007 (-0.009, -0.005) ^**^ | -0.043 (-0.068, -0.017) ^**^ | -0.003 (-0.005, -0.001) ^**^ | -0.035 (-0.064, -0.005) ^*^ | -0.001 (-0.005, 0.003) | -0.081 (-0.094, -0.067) ^**^ | -0.008 (-0.009, -0.007) ^**^ |
| * p < 0.05. ** p < 0.05/3 (Bonferroni correction). CHARLS=China Health and Retirement Longitudinal Study. HRS=Health and Retirement Study. ELSA=English Longitudinal Study of Ageing. PP=pulse pressure. models were adjusted for baseline age, sex, other modifiable factors including early life factor (education), midlife factors (hearing loss, depression, smoking, drinking, diabetes, hypertension, obesity, and physical inactivity), and late life factors (marital status and visual loss). | | | | | | | | |

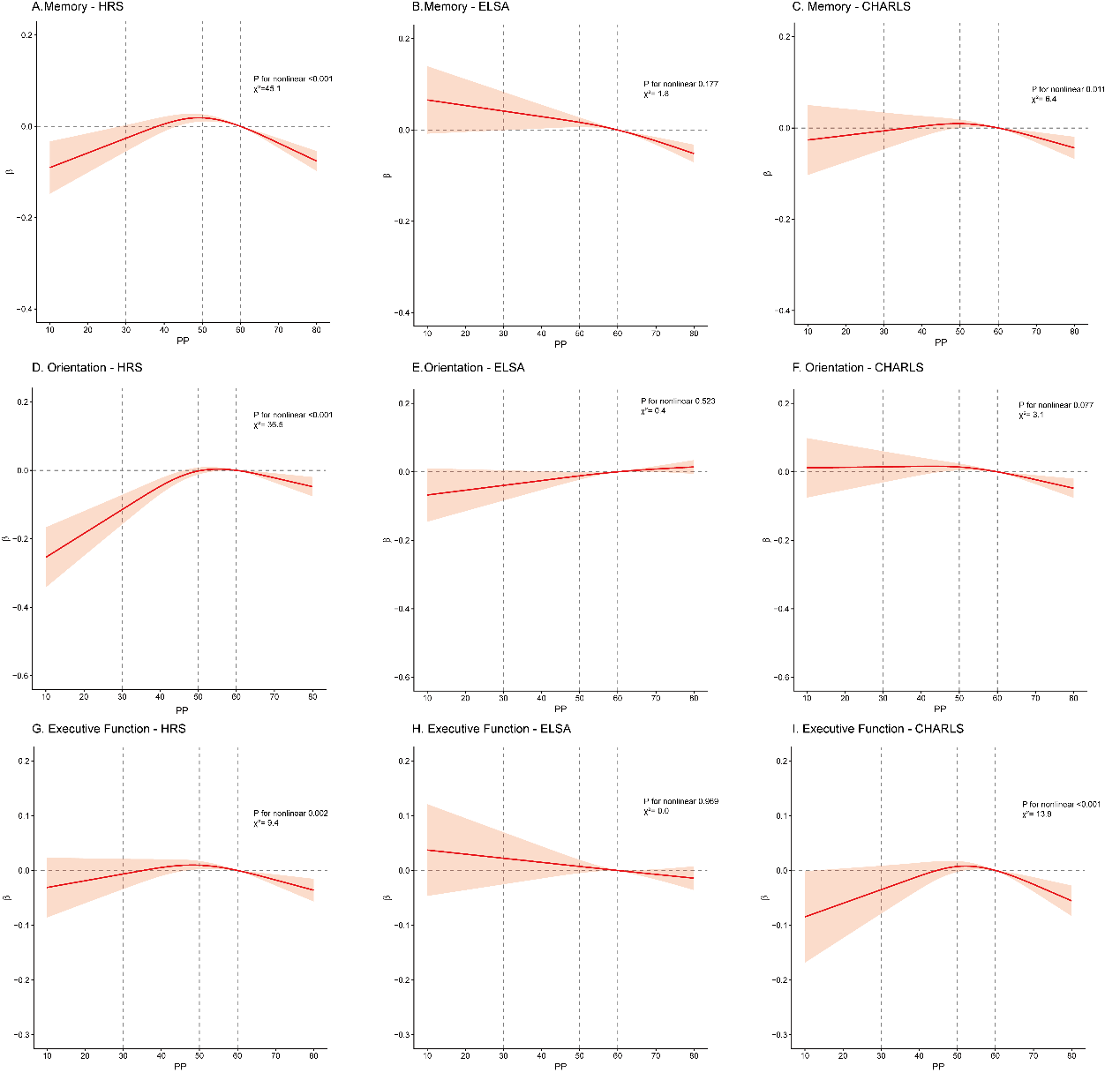

**Figure S1: Nonlinear associations of PP with memory (A-C), orientation (D-F), and execution (G-I) in HRS (A, D, G), ELSA (B, E, H), and CHARLS (C, F, I)**

P values represents the overall test for nonlinearity estimated from linear mixed-effects models. Reference: PP =60 mmHg. Nonlinear effects of baseline PP were modelled using a restricted cubic spline with three knots. Models were adjusted for age, sex, mean arterial pressure, early life factor (education), midlife factors (hearing loss, depression, smoking, drinking, diabetes, hypertension, obesity, and physical inactivity), and late life factors (marital status and visual loss). CHARLS=China Health and Retirement Longitudinal Study. ELSA=English Longitudinal Study of Ageing. HRS=Health and Retirement Study. PP=pulse pressure.

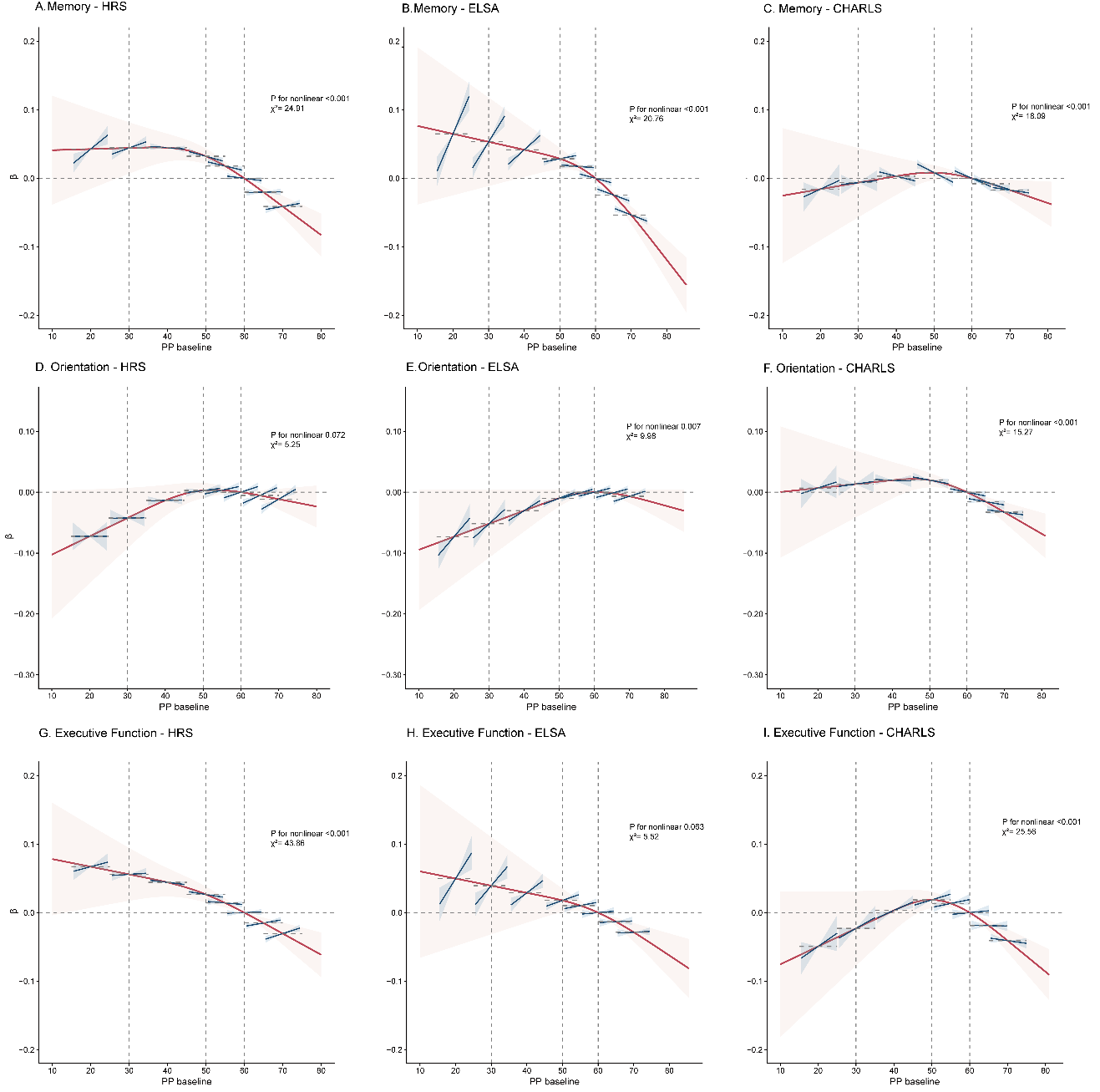

**Figure S2: Nonlinear associations of PP baseline and change with memory (A-C), orientation (D-F), and execution (G-I) in HRS (A, D, G), ELSA (B, E, H), and CHARLS (C, F, I)**

Models were specified as: Cognition ~ Spline (PP baseline) * PP change + Age + Covariates + (1|Participant). P values represents the overall test for nonlinearity estimated from linear mixed-effects models. The red line represents the coefficients of PP baseline (reference PP baseline=60 mmHg), whereas the blue line represents the coefficients of PP change at the corresponding level of PP baseline (reference PP change=0 mmHg). To improve clarity and reduce the influence of nonlinearity in PP change, PP change is presented within the range of −5 to +5. Nonlinear effects of baseline PP were modelled using a restricted cubic spline with three knots. Covariates included sex, mean arterial pressure, early life factor (education), midlife factors (hearing loss, depression, smoking, drinking, diabetes, hypertension, obesity, and physical inactivity), and late life factors (marital status and visual loss). CHARLS=China Health and Retirement Longitudinal Study. ELSA=English Longitudinal Study of Ageing. HRS=Health and Retirement Study. PP=pulse pressure.

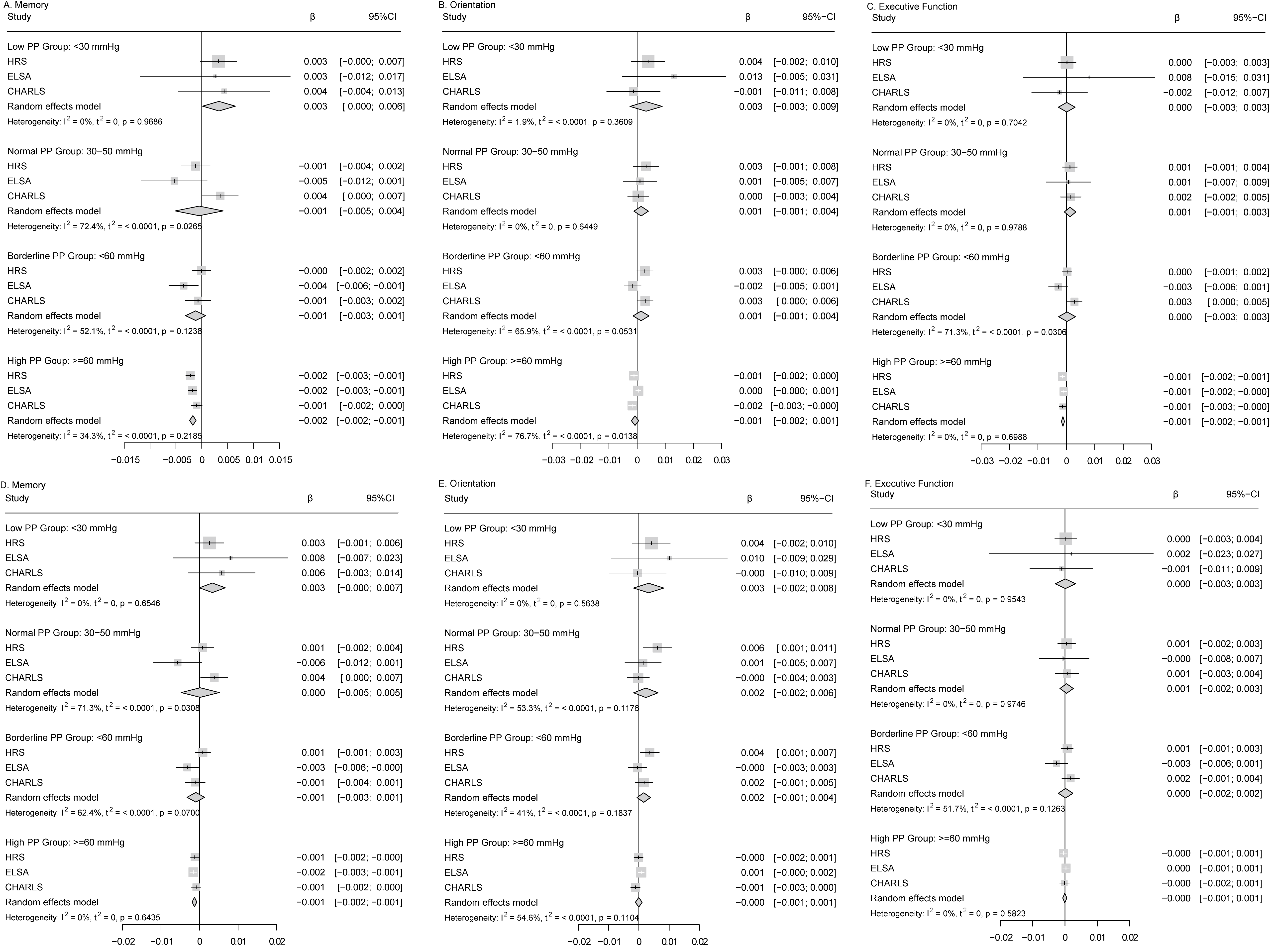

**Figure S3: Associations of PP with Memory (A, D), Orientation (B, E), and Executive function (C, F) stratified by PP threshold groups**

Coefficients of PP were presented. A-C. *Cognition ~ PP + Age + (1|Participant).* D-F. All models were additionally adjusted for sex, MAP, and modifiable factors across early, mid, and late life.

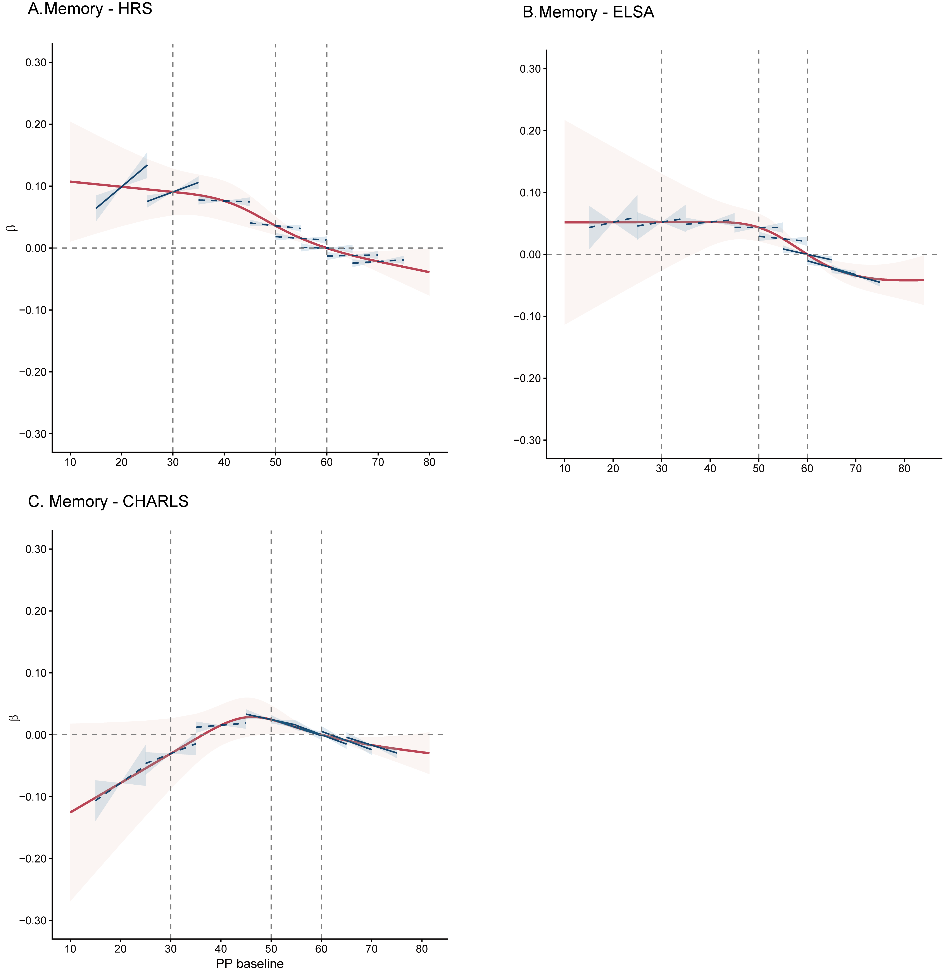

**Figure S4: Associations of PP baseline and change with memory**

Models were specified as: Memory ~ Spline (PP baseline) * PP change + Spline (Age baseline) + Time + Covariates + (1|Participant). P values represents the overall test for nonlinearity estimated from linear mixed-effects models. The red line represents the coefficients of PP baseline (reference PP baseline=60 mmHg), whereas the blue line represents the coefficients of PP change at the corresponding level of PP baseline (reference PP change=0 mmHg). To improve clarity and reduce the influence of nonlinearity in PP change, PP change is presented within the range of −5 to +5. Nonlinear effects of baseline PP were modelled using a restricted cubic spline with three knots. Models were based on Model 3.2 and further adjusted for sex, mean arterial pressure, early life factor (education), midlife factors (hearing loss, depression, smoking, drinking, diabetes, hypertension, obesity, and physical inactivity), and late life factors (marital status and visual loss). CHARLS=China Health and Retirement Longitudinal Study. ELSA=English Longitudinal Study of Ageing. HRS=Health and Retirement Study. PP=pulse pressure.
